# Air pollution and emotional behavior in adolescents across the U.S.

**DOI:** 10.1101/2023.04.19.23288834

**Authors:** Claire E. Campbell, Devyn L. Cotter, Katherine L. Bottenhorn, Elisabeth Burnor, Hedyeh Ahmadi, W. James Gauderman, Carlos Cardenas-Iniguez, Daniel Hackman, Rob McConnell, Kiros Berhane, Joel Schwartz, Jiu-Chiuan Chen, Megan M. Herting

## Abstract

Recent studies have linked air pollution to increased risk for behavioral problems during development, albeit with inconsistent findings. Additional longitudinal studies are needed that consider how emotional behaviors may be affected when exposure coincides with the transition to adolescence – a vulnerable time for developing mental health difficulties. This study examines how annual average PM_2.5_ and NO_2_ exposure at ages 9-10 years relates to internalizing and externalizing behaviors over a 2-year follow-up period in a large, nationwide U.S. sample of participants from the Adolescent Brain Cognitive Development (ABCD) Study®. Air pollution exposure was estimated based on the residential address of each participant using an ensemble-based modeling approach. Caregivers answered questions from the Child Behavior Checklist (CBCL) at baseline and annually for two follow-up sessions for a total of 3 waves of data; from the CBCL we obtained scores on internalizing and externalizing problems plus 5 syndrome scales (anxious/depressed, withdrawn/depressed, rule-breaking behavior, aggressive behavior, and attention problems). Zero-inflated negative binomial models were used to examine both the main effect of age as well as the interaction of age with each pollutant on behavior while adjusting for various socioeconomic and demographic characteristics. Overall, the pollution effects moderated the main effects of age with higher levels of PM_2.5_ and NO_2_ leading to an even greater likelihood of having no behavioral problems (i.e., score of zero) with age over time, as well as fewer problems when problems are present as the child ages. Albeit this was on the order equal to or less than a 1-point change. Thus, one year of annual exposure at 9-10 years is linked with very small change in emotional behaviors in early adolescence, which may be of little clinical relevance.

## 1. Introduction

Mental health conditions remain a global health challenge for all age groups (World Health Organization, 2018), but the risk for onset of psychopathology is highest in childhood and adolescence. Both internalizing and externalizing symptoms typically emerge during adolescence (Achenbach et al., 1991). Moreover, up to approximately 20% of children and adolescents are affected by mental health problems worldwide (Polanczyk et al., 2015) with half of all lifetime mental health conditions diagnosed by age 14 years (Kessler et al., 2005). In order to reduce societal costs and improve quality of life for affected individuals, research on modifiable risk and resilience factors holds the promise to potentially uncover new avenues for early prevention and intervention.

Recent evidence indicates that outdoor air pollution may contribute to increased risk for mental health conditions (Braithwaite et al., 2019; Zundel et al., 2022). A growing body of literature has associated ambient exposure to fine particulate matter (PM_2.5_) and nitrogen dioxide (NO_2_) with mental health disorder outcomes in children, adolescents, and adults, including symptoms of anxiety, depression, and aggression in children, as well as increased risk for attention deficit and hyperactivity disorder (ADHD) and delinquency problems (Forns et al., 2016; Margolis et al., 2016; Newman et al., 2013; Perera et al., 2014, 2018; Thygesen et al., 2020; Yorifuji et al., 2016, 2017). However, recent comprehensive reviews of how air pollution relates to anxiety and depression (Zundel et al., 2022) and on attention problems (Myhre et al., 2018) both highlight a number of inconsistencies and important knowledge gaps in the broader air pollution and mental health literature. For example, while the majority of studies found positive associations between air pollution exposure and anxiety and depression, 25% of studies did not find associations or reported mixed effects and fewer than 10% of all air pollution and internalizing behavior studies examined air pollution exposure during the susceptible window of childhood and adolescence. Even within the developmental literature, there are mixed results: some studies suggest prenatal through adolescent exposure is linked to more internalizing problems (Margolis et al., 2016; Brokamp et al., 2019; Brunst et al., 2019; B. Fan et al., 2019; Rasnick et al., 2021; Yolton et al., 2019), where others have failed to find an association (Jorcano et al., 2019; Zhao et al., 2019). One potential limitation of many of these studies is a large age range over which links between exposure and mental health are assessed, which may fail to capture age-specific or nonlinear effects.

Importantly, most previous studies have also been limited to cross-sectional assessment of mental health outcomes and/or are limited in terms of both the geographic and sociodemographic diversity of their study sample. Discrepancies in results may also be due in part to both differences in the window of exposure, as well as the timing of the behavioral evaluation, especially given the known developmental patterns in symptom onset. For example, a recent study of 8 European cohorts found that neither prenatal nor early life exposure was related to cross-sectional outcomes of depression, anxiety, or aggression behavior using the Child Behavior Checklist (CBCL) or Strengths and Difficulties Questionnaire (SDQ) when assessed in mid-to-late childhood (Jorcano et al., 2019). This highlights an additional drawback of large, multi-cohort studies: different instruments are often used to measure similar behaviors, and often only a narrow range of internalizing and externalizing behaviors are addressed. On the other hand, Roberts and colleagues (2019) found that while exposure to higher levels of PM_2.5_ and NO_2_ at age 12 years was not associated with concurrent mental health conditions, it did successfully predict a 1.5 fold increased risk for developing major depressive disorder when assessed longitudinally at 18 years of age. Similarly, Reuben and colleagues (2021) found that exposure to nitrous oxides (NO_X_) in childhood predicted later onset of internalizing, externalizing, and thought disorder symptoms at age 18 years. The latter two studies suggest that exposure during adolescence may ultimately predispose an individual to later develop mental health conditions, further emphasizing the importance in assessing emotional and behavioral problems longitudinally. Furthermore, these studies were limited in that one of them was based on a smaller sample size and both were limited geographically to England and/or Wales. Thus, large and geographically diverse samples with repeated assessments of mental health are necessary to improve the potential generalizability as to whether exposure during the transition to adolescence may contribute to increased risk in the development of emotional behavioral problems.

Beyond the considerations of timing of the exposure and longitudinal assessment of the outcome, questions also remain as to the potential health effects of air pollution below the current air quality standards. That is, despite significant declines in air pollution, recent epidemiological studies continue to find links between adverse effects and levels of exposure well below the Environmental Protection Agency (EPA)’s specified annual averages for PM_2.5_ (<= 12 μ*g/m^3^*) and NO_2_ (<=53 *parts per billion; ppb*) (Dominici et al., 2019). This emerging body of research showing that health effects, including cardiovascular and respiratory disease-related morbidity and mortality, can be observed at much lower concentrations of exposure suggests that no observable threshold may be considered “safe” (for review, see Papadogeorgou et al., 2019). As such, the World Health Organization (WHO) updated their regulatory guidelines on ambient air quality in September 2021, recommending that annual averages of PM_2.5_ and NO_2_ not exceed 5 μ*g/m^3^*and 10 μ*g/m^3^* (equivalent to 18.8 *ppb*) respectively (*WHO Global Air Quality Guidelines*, 2021). Given exposure levels captured in previous studies associating ambient air pollution and behavioral problems have largely fallen above EPA standards, further research is necessary to investigate to what degree low-levels of exposure may influence emotional behavior in today’s youth.

To address these gaps in knowledge, we leveraged the large (N=11,876), nationwide, and socio-demographically diverse Adolescent Brain Cognitive Development (ABCD) Study^®^ cohort (Jernigan et al., 2018) to examine whether air pollution exposure at ages 9-10 years may relate to longitudinal changes in behavioral and emotional problems over a 2-year follow-up period. This targeted age range allows us to isolate associations during a crucial time of development: the transition to adolescence and onset of puberty. The ABCD Study^®^ comprises 21 study sites across the United States and implements an identical protocol for recruitment and data collection of all participants (Garavan et al., 2018; Lisdahl et al., 2018). The richness of the ABCD dataset also allows us to examine and adjust for vital confounders to improve the rigor and robustness of our findings. Since its sample is geographically diverse and participants were enrolled between October 2016 and 2018, the ABCD dataset can address links between air pollution and mental health in those regularly exposed to concentrations largely below EPA standards. Given the previous findings (Margolis et al., 2016; Brokamp et al., 2019; Brunst et al., 2019; B. Fan et al., 2019; Rasnick et al., 2021; Yolton et al., 2019), we *a priori* chose to examine internalizing and externalizing summary scores from the Child Behavior Checklist (CBCL), as well as distinct internalizing syndrome subscales of anxious/depressed and withdrawn/depressed, and externalizing syndrome subscales of rule-breaking and aggressive behavior, and the independent subscale of attention, thus addressing a wide range of internalizing and externalizing behaviors. We hypothesized that higher exposure levels during late childhood would predict more emotional problems over a 2 year follow-up period.

## 2. Materials and methods

### 2.1 Study Design and Participants

The current study utilized data from the larger ongoing nationwide ABCD Study^®^ (NDA 4.0 data release 2021, https://abcdstudy.org/scientists/data-sharing/), which enrolled over 11,876 9- and 10-year-old participants across the USA from 2016-2018 (Garavan et al., 2018; Jernigan et al., 2018; Volkow et al., 2018). The 21 study sites obtained approval from their local Institutional Review Board (IRB) and a centralized IRB approval was obtained from the University of California, San Diego. Written informed consent was provided by each child’s parent or legal guardian (hereafter, “caregiver”); each child provided verbal assent. All ethical regulations were complied with during data collection and analysis. Primary inclusion criteria for ABCD Study participants included age (9.0 to 10.99 years at baseline visit), fluency in English, and the ability to complete the baseline visit. For the current analysis, we utilized data from the first 3 waves of annual data collection, with the additional inclusion criteria of having a valid primary residential address at baseline for all subjects. Given the distribution of the outcome data and analytic approach required for hypothesis testing (see section 2.5 below), complete cases were required for each wave of data collection. We also selected data collected before March 1, 2020, to avoid any potential confounding effects of stress on mental health outcomes related to the COVID-19 pandemic (Hamatani et al., 2022; Kiss et al., 2022; Yip et al., 2022). Lastly, we randomly selected one subject per family to reduce the hierarchical structure of our data from 4 levels (time point, subject, family, site) to 3 levels (time point, subject, site). This resulted in a final sample of 9,271 participants (**Table 1**). A comparison of the overall ABCD cohort with our final analytical sample at baseline, 1-year, and 2-year follow-up can be found in **Supplemental Table 1**, **Supplemental Table 2**, and **Supplemental Table 3**, respectively. All variable names used in the following analyses are documented in the **Supplemental Table 8**.

**Table 1.** Demographic composition of the final sample across three waves of data collection.

|  | Baseline | 1-year follow-up | 2-year follow-up |
| --- | --- | --- | --- |
| N | 9271 | 8759 | 5827 |
| Sex assigned at birth |  |  |  |
| Female | 4413 (47.6%) | 4154 (47.4%) | 2747 (47.1%) |
| Male | 4858 (52.4%) | 4605 (52.6%) | 3080 (52.9%) |
| Age at data collection |  |  |  |
| Mean (SD) | 9.91 (0.62) | 10.91 (0.63) | 11.92 (0.64) |
| Range | 8.92 - 11.08 | 9.75 - 12.42 | 10.58 - 13.67 |
| Race/ethnicity |  |  |  |
| Black | 1363 (14.7%) | 1221 (13.9%) | 678 (11.6%) |
| Hispanic | 1958 (21.1%) | 1800 (20.6%) | 1198 (20.6%) |
| Other | 1191 (12.8%) | 1126 (12.9%) | 715 (12.3%) |
| White | 4759 (51.3%) | 4612 (52.7%) | 3236 (55.5%) |
| Caregiver education |  |  |  |
| < HS Diploma | 460 (5.0%) | 405 (4.6%) | 257 (4.4%) |
| HS Diploma/GED | 899 (9.7%) | 800 (9.1%) | 460 (7.9%) |
| Some College | 2423 (26.1%) | 2247 (25.7%) | 1466 (25.2%) |
| Bachelor | 2310 (24.9%) | 2217 (25.3%) | 1551 (26.6%) |
| Post Graduate Degree | 3179 (34.3%) | 3090 (35.3%) | 2093 (35.9%) |
| Caregiver employment |  |  |  |
| Employed | 6444 (69.5%) | 6146 (70.2%) | 4139 (71.0%) |
| Stay at Home Parent | 1612 (17.4%) | 1516 (17.3%) | 1002 (17.2%) |
| Unemployed | 539 (5.8%) | 481 (5.5%) | 299 (5.1%) |
| Other | 676 (7.3%) | 616 (7.0%) | 387 (6.6%) |
| Neighborhood safety |  |  |  |
| Mean (SD) | 3.873 (0.976) | 3.884 (0.971) | 3.915 (0.948) |
| Range | 1.000 - 5.000 | 1.000 - 5.000 | 1.000 - 5.000 |
| Household income |  |  |  |
| <\$50k | 2564 (27.7%) | 2335 (26.7%) | 1597 (27.4%) |
| ≥\$50K & <\$100K | 2419 (26.1%) | 2312 (26.4%) | 1597 (27.4%) |
| ≥\$100K | 3514 (37.9%) | 3418 (39.0%) | 2307 (39.6%) |
| Don't know or refuse | 774 (8.3%) | 694 (7.9%) | 429 (7.4%) |

### 2.2 Estimation of Annual Air Pollution Exposure

Details regarding the collection of residential addresses and linkage to ambient PM_2.5_ and NO_2_have been previously published in detail by Fan and colleagues (2021). Briefly, daily pollutant estimates were derived at a 1-km^2^ resolution using hybrid spatiotemporal models that combine satellite-based aerosol optical depth models, land-use regression, and chemical transport models (Di et al., 2019, 2020). The cross-validation of these models with EPA monitored levels across the U.S. were found to perform well, with R^2^ Root Mean Square Error of 0.89 for PM_2.5_ annual averages and 0.84 for NO_2_ annual averages (Di et al., 2019, 2020). These daily estimates were then averaged over the 2016 calendar year, when the children were aged 9-10 years-of-age (baseline age) and assigned to the geocoded primary residential address at the baseline study visit. PM_2.5_ is reported in micrograms per meter cubed (*µg/m^3^*) and NO_2_ is reported in parts per billion (*ppb*).

### 2.3 Emotional Behavior

At baseline and each follow-up annual visit, the participant’s caregiver was asked to report on the child’s emotional behavior over the 6 months prior to each study visit using the Child Behavioral Checklist (CBCL) (Achenbach, 2009; Achenbach & Rescorla, 2001). The CBCL has 120 different items that each caregiver answers about their child (e.g., “Show little interest in things around him/her”) using a 3 point Likert-type scale (0 = Not True, 1 = Somewhat or Sometimes True, 2 = Very True or Often True). These answers are then used to calculate summary scores of internalizing and externalizing behavior. Based on the prior air pollution and behavioral literature (for review see Zundel et al., 2022a; Margolis et al., 2016; Brokamp et al., 2019; Brunst et al., 2019; B. Fan et al., 2019; Rasnick et al., 2021; Yolton et al., 2019), we also chose to examine five additional syndrome subscale scores: anxious/depressed, withdrawn/depressed, rule-breaking behavior, aggressive behavior, and attention problems. Anxious/depressed and withdrawn/depressed subscales fall within the internalizing score, rule-breaking and aggressive behavior subscales fall within the externalizing score, and attention is an independent subscale. Each raw score is a whole number with higher integers indicating increased problem or emotional behaviors. Importantly, the CBCL measures show good test-retest reliability (Pearson’s correlations mean = 0.9 (min=0.82, max=0.94) and internal consistency was stable over a 12- and 24-month period (Pearson’s correlation 12-month mean = 0.74m, 24-month mean = 0.70) (Achenbach & Rescorla, 2001). For the current study we utilized the CBCL subscale raw scores to examine the influence of change over time. The raw scores have different ranges by subscale: internalizing [0,64], externalizing [0,70], anxious/depressed [0,26], withdrawn/depressed [0,16], rule-breaking [0,34], aggressive [0,36], and attention [0,20], with higher values indicative of greater problems.

### 2.4 Confounders and Covariates

We have selected potential confounders based on both prior knowledge and current theories in environmental epidemiology. We have identified a list of variables that are likely related to air pollution exposure, all of which were reported by the child’s caregiver using the PhenX Toolkit (Echeverria et al., 2004; Mujahid et al., 2007). This list includes child’s sex, race/ethnicity, highest caregiver educational attainment, caregiver’s employment status, perceived neighborhood safety, and the combined total annual household income. In the ABCD Study, the race/ethnicity variable also includes ‘Asian’ as a category, but due to a low number of individuals who fall within this category, ‘Asian’ had to be merged with ‘Other’. Each of these variables’ baseline values were used in the model, to align with the timing of the available ambient air pollution estimates. To account for potential confounding of co-exposure, we also included the other air pollutant as an additional variable (i.e., when examining the influence of PM_2.5_-by-age on CBCL outcomes, NO_2_ is added to the model, and vice versa). Importantly, multicollinearity was not an issue in adjusting for the other pollutant in the model as the Pearson correlation coefficient between the baseline annual pollutant concentrations of PM_2.5_ and NO_2_ across all sites was low (*r* = 0.22).

### 2.5 Analyses

All statistical analyses were implemented in R (version 4.1.2) (R Core Team, 2021). Initial descriptive and exploratory analysis were conducted to check all data for potential errors and outliers and to assess variable distributions required to satisfy modeling assumptions and to understand correlations. To examine how age and annual PM_2.5_ and NO_2_ relate to emotional behaviors at ages 9-10 years, as well as over a 1- and 2-year follow-up, we used a multilevel (i.e., mixed effects) modeling approach. Since CBCL outcomes are naturally zero-inflated (**Supplemental Figure 1**) and thus over-dispersed (over-dispersion quotient ranges from 2.84-17.29), using a linear mixed-effects model can lead to artificial inflation of the coefficients’ significance (LAND et al., 1996; Stroup, 2016; Swartout et al., 2015). Previous papers examining CBCL outcomes and air pollution have examined the CBCL t-scores (Perera et al., 2011, 2012; Jorcano et al., 2019) which is useful given their normal distributions, but does not allow for the investigation of an interaction with age. To address these concerns of zero-inflation, we treated the CBCL outcomes as count data and employed a zero-inflated negative binomial (ZINB) model, which adds an extra parameter that accounts for over-dispersion (Xu et al., 2017). The *glmm.zinb()* function was used within the NBZIMM package (version 1.0) (https://github.com/nyiuab/NBZIMM); a manuscript detailing the development of this package was also published (Zhang & Yi, 2020). This modeling approach has been used in numerous studies with zero-inflated health data (Preisser et al., 2016; Sheu et al., 2004), and specifically when examining mental health outcomes (Kumagai et al., 2021; Vyas et al., 2020). To implement this model, complete data across timepoints (i.e., no missing values for each subject at each wave of data collection) is required, so listwise deletion was used to remove incomplete data by wave of data collection. This means that, following our initial cleaning steps, creating a complete dataset across timepoints led to 6%, 5%, and 3% of missing data for the baseline, 1-year, and 2-year follow-up visits, respectively. Bennet (2001) states that greater than 10% missingness could lead to bias within the statistical analysis and prior published literature suggest 5% (on average) missingness is negligible (Jakobsen et al., 2017; Schafer, 1999). Therefore, given the limited amount of missing data, we chose not to perform multiple imputation.

The ZINB model combines two models: 1) zero-inflated, model similar to a logistic regression, that evaluates the likelihood of being in the certain-zero as compared to the non-zero category, and 2) count model assuming a negative binomial distribution that evaluates the non-zero CBCL subscale scores. Initially, age-only models were performed to establish the age relationship associated with each CBCL outcome. These models investigated the main effect of age on each CBCL outcome controlling for necessary covariates in both the zero-inflated and count portions of the model: child’s sex, race/ethnicity, highest caregiver educational attainment, caregiver’s employment status, perceived neighborhood safety, and the combined total annual household income. For the random effects within the zero-inflated portion of the model, we only included ABCD site since subjects within the certain-zero group had extremely low intraclass correlation coefficients (ICC) for all CBCL outcomes examined (ranging from 0.070-0.107; implying the data were not strongly clustered by subject). For the random effects within the count model, subject was nested within a random effect of site to account for the within-subject similarities over time in those with non-zero data (ICCs ranging from 0.506-0.710; this medium-high ICC implies a clustering structure of subject within the non-zero data). To investigate the effect of air pollution on CBCL outcomes, we added in an interaction between age and each air pollutant (PM_2.5_ or NO_2_). For both the zero-inflated model and the count model we utilized the fixed effects of each pollutant (PM_2.5_ or NO_2_), age, pollutant-by-age, while adjusting for potential confounders as mentioned above; each pollution-by-age model also corrected for the other pollutant (e.g., for the PM_2.5_-by-age model, NO_2_ was added as a confounder in addition to the previously mentioned covariates, and vice versa for the NO_2_-by-age model). For the random effects within the zero-inflated portion of the model, we again only included ABCD site since subjects within the certain-zero group had extremely low ICC for all CBCL outcomes; for the random effects within the count model, subject was nested within a random effect of site to account for the within-subject similarities over time in those with non-zero data. PM_2.5_ and NO_2_ were centered to the levels recommended by the WHO, 5 µg/m^3^ and 5.33 ppb, respectively, and age was centered at 9 years (the youngest integer age in our cohort). For models where the interaction term between pollution and age was not significant for both the zero-inflated and count portion of the model, the interaction term was dropped, and the model was run to examine the main effect of pollution. To avoid type-1 errors, all *p*-values of interest were corrected for multiple comparisons across the same model type using the false-discovery rate of 5% by utilizing the Benjamin-Hochberg procedure (Benjamini & Hochberg, 1995), which has been used previously with a ZINB modeling approach (Subramaniyam et al., 2019). All model assumptions post analyses were also conducted based on prior published methodology (Cameron & Trivedi, 2013; Garay et al., 2011; Hilbe, 2011). In terms of interpreting our PM_2.5_ results, we focused on displaying the predictions of the EPA annual daily standard (PM_2.5_ = 12 µg/m^3^) as compared to the WHO’s recommended target level of 5 µg/m^3^. For NO_2_, our sample’s exposure levels were much less than the 53 ppb previously set by EPA in 1971 (US EPA, 2016). Thus for NO_2_ we focused on comparing predictions at 26.1 ppb, based on the 90th percentile of our sample, as compared to the WHO recommended 5.33 ppb. Lastly, given that very large sample sizes tend to identify very small differences as significant, we were sure to also interpret our results in context of effect sizes in order to assess if results were likely to be clinically significant as defined by Jacobson & Truax (1991), which requires not only statistical significance, but also a change either in the range of the “dysfunctional population” or “within the range of the functional population”.

## 3. Results

Descriptives of our analytical dataset separated by baseline, 1-year, and 2-year follow-up can be found in **Table 1**. The mean for PM_2.5_ for the total current sample was 7.706 µg/m^3^ (range=1.722,15.902 SD=1.571) and for NO_2_ it was 18.595 ppb (range=0.729,37.940; SD=5.571), which on average falls significantly below the EPA standards (p’s<0.0001) of 12 µg/m^3^ and 53 ppb, respectively. Furthermore, descriptives of CBCL outcomes across waves of data collection are presented in **Supplemental Table 4**.

### 3.1 Internalizing Behavior

For internalizing behavior, we found a significant main effect of age for the zero-inflated portion of the model, demonstrating a 45% increase in the probability of having zero internalizing problems (i.e., obtaining a true-zero) with increasing age (**Supplemental Figure 2)**. Additionally, we found a significant interaction between PM_2.5_ and age for both the zero-inflated and count portions of the model (**Figure 1**, **Supplemental Table 5**). We found that for a PM_2.5_ concentration of 5 µg/m^3^ (i.e., the WHO standard), there was a 13% decrease in the probability of having zero internalizing problems (i.e., obtaining a true-zero) as age increases from 9 to 12 years-of-age, whereas at a higher PM_2.5_ concentration of 12 µg/m^3^ there was a 190% increase in the probability of having no internalizing problems (i.e., true-zero) from 9 to 12 years-of-age. When examining the count model, there was a 9% increase versus a 13% decrease in internalizing scores from ages 9 to 12 years at PM_2.5_ of 5 µg/m^3^ and 12 µg/m^3^, respectively. A similar trend was seen for the influence of NO_2_-by-age on internalizing behavior (**Figure 1, Supplemental Table 7**). For 5.33 ppb (i.e., WHO recommendation) the probability of having no internalizing problems (i.e., obtaining a true-zero) from 9 to 12 years-of-age decreased by 12%, whereas the probability of having no internalizing problems increases by 106% from 9 to 12 years-of-age at 26.1 ppb of NO_2_ (i.e., 90^th^ percentile of sample); in the count model, there was a 13% increase versus a 6% decrease in internalizing scores from 9 to 12 years-of-age at 5.33 ppb and 26.1 ppb NO_2_ concentrations, respectively. Although, it is important to note these detected changes in probability still average below 5% probability of no internalizing problems (i.e., true-zero score) as well as only related to less than a 1-point change in the non-zero CBCL internalizing scores themselves.

**Figure 1.**
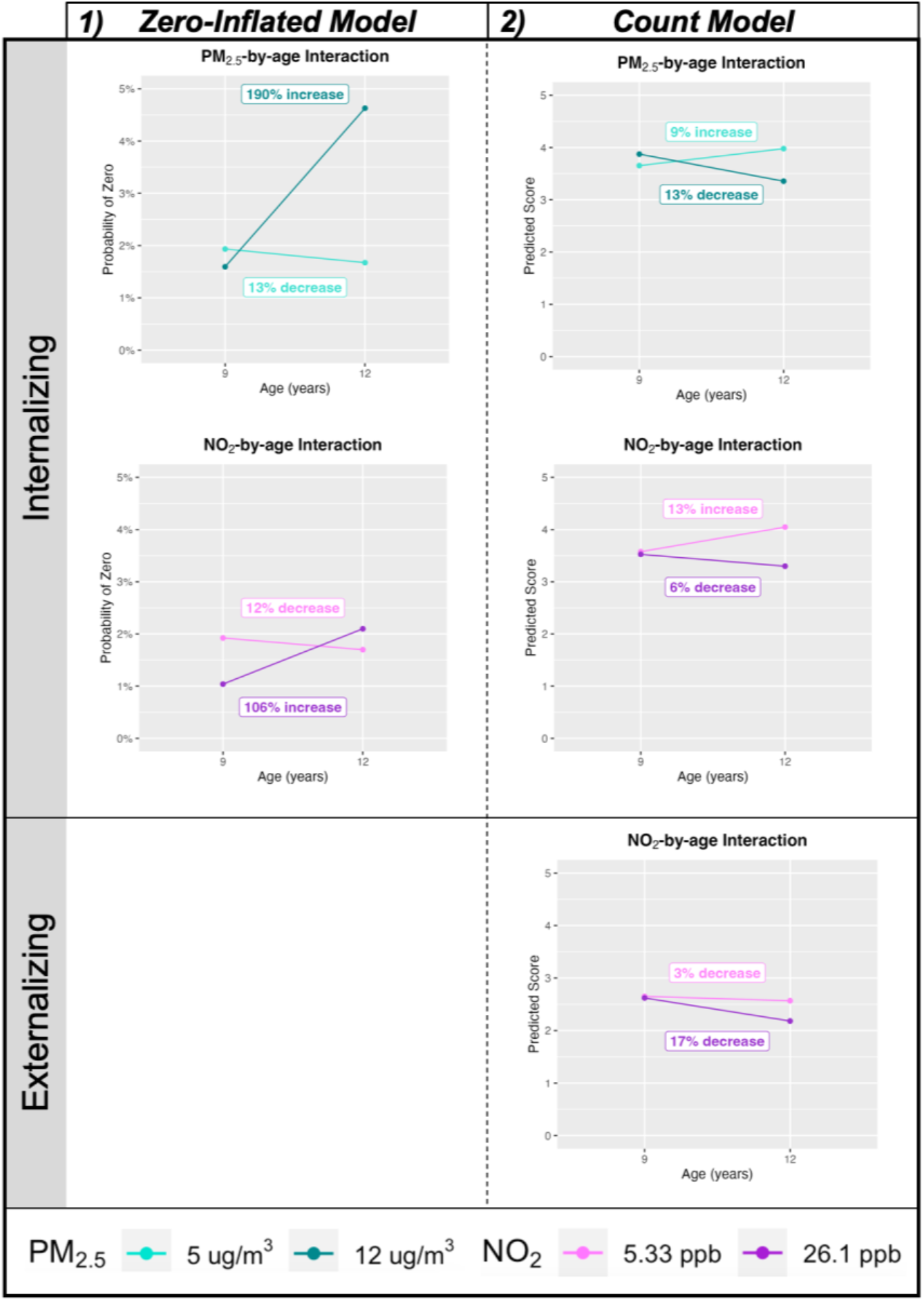
Significant results for internalizing and externalizing behavior. NO_2_ is set to the WHO standard (5.33 ppd) for the PM_2.5_-by-age models and PM_2.5_ is set to the WHO standard (5 µg/m^3^) for the NO_2_-by-age models. ***Column 1)*** Displays the estimated probability of being in the absolute zero category as compared to the non-zero category (i.e., any value for CBCL scores). ***Column 2)*** Displays the estimated CBCL score for subjects whose scores were in the non-zero category. Numerous results are presented which include: ***a)* PM_2.5_-by-age interaction** which displays differences in 9 and 12 years-of-age for the WHO recommended PM_2.5_ levels - 5 µg/m^3^ (light blue) - versus the EPA’s - 12 µg/m^3^ (dark blue); ***b)* NO_2_-by- age interaction** which displays differences in 9 and 12 years-of-age for the WHO recommended NO_2_ levels - 5.33 ppd (light purple) - versus the 90^th^ percentile NO_2_ level in our sample - 26.1 ppd (dark purple) (The EPA level is 53 ppd which is outside our sample range). All graphs display percent change with age. All covariates held constant at the largest N category (sex = “male”, race/ethnicity = ‘White’, caregiver education = ‘Post Graduate Degree’, caregiver employment = “Employed”, and household income = “≥$100K”), and mean for neighborhood safety (𝑥 = 3.88)

### 3.2 Externalizing Behavior

For externalizing behavior, we found a significant main effect of age for the zero-inflated portion of the model, demonstrating a 88% increase in the probability of having zero externalizing problems (i.e., obtaining a true-zero) with increasing age, as well as for the count portion of the model, showing a 12% decrease in problems with increasing age (**Supplemental Figure 2)**. Additionally, there was no significant interaction between age and PM_2.5_ on either the zero-inflated or the count portions of the model, but there was a main effect of exposure within the zero-inflated model once the interaction term was dropped (**Figure 1**, **Supplemental Table 6**). At PM_2.5_ concentration of 5 µg/m^3^, there was a 1.5% probability of having no externalizing problems (i.e., a true-zero), whereas at a PM_2.5_ concentration of 12 µg/m^3^ there was a 60% increase in the probability of having no externalizing problems. There was no significant main effect of PM_2.5_ on non-zero externalizing behavior scores. For the NO_2_-by-age model, the zero-inflated portion of the model was not significant, meaning there was no difference between NO_2_ exposure levels and the probability of having no externalizing problems across age; however, the non-zero portion of the model was significant. At 5.3 ppb, externalizing scores decreased by 3% from 9 to 12 years-of-age, while at the NO_2_ concentration of 26.1 ppb a 17% decrease in externalizing scores was seen from 9 to 12 years-of-age (**Figure 1**). Though, again, the magnitude of these changes in predicted CBCL non-zero scores were less than a 1-point change.

### 3.3 Subscales

#### 3.3.1 Anxious/Depressed and Withdrawn Depressed

These subscales both fall within the internalizing score, therefore, unsurprisingly, they showed very similar trends to the overall internalizing score (**Figure 2** **and Supplemental Table 5**). The anxious/depressed subscale showed a significant main effect of age for the zero-inflated portion of the model, demonstrating a 185% increase in the probability of having zero problems (i.e., obtaining a true-zero) with increasing age, as well as for the count portion of the model, showing a 8% decrease in problems with increasing age (**Supplemental Figure 3).** The withdrawn/depressed subscale showed a significant main effect of age for the zero-inflated portion of the model, demonstrating a 45% decrease in the probability of having zero problems (i.e., obtaining a true-zero) with increasing age, as well as for the count portion of the model, showing a 34% increase in problems with increasing age (**Supplemental Figure 3).** Additionally, there was a significant interaction for PM_2.5_-by-age in the zero-inflated portion of the model, with a larger relative increase in the probability of having no problems (i.e., true zero) from 9 to 12 years-of-age at a PM_2.5_ concentration of 12 µg/m^3^ of PM_2.5_ as compared to 5 µg/m^3^, which the withdrawn/depressed subscale was non-significant. For the non-zero count portion of the model, significant interactions were seen between PM_2.5_ and age for both the anxious/depressed and withdrawn/depressed, with a 9% increase in anxious/depressed and 46% increase in withdrawn/depressed from ages 9-12 years at PM_2.5_ concentrations of 5 µg/m^3^, but a decrease of 13% and increase of 19% at a PM_2.5_ concentration of 12 µg/m^3^, respectively. Similar patterns were also seen for significant NO_2_-by-age effects on both the zero-inflated and count portions of the anxious/depressed and withdrawn/depressed subscales. Specifically, at NO_2_ concentrations of 26.1 ppb, there was a greater probability of reporting no problems (i.e., true-zero) as well as relatively less anxious/depressed or depressed/withdrawn scores at 12 years-of-age as compared to NO_2_ concentrations of 5.3 ppb. However, the magnitude of probabilities again was within a 5% probability range within the zero-inflated model and less than a 1-point change in the predicted scores.

**Figure 2.**
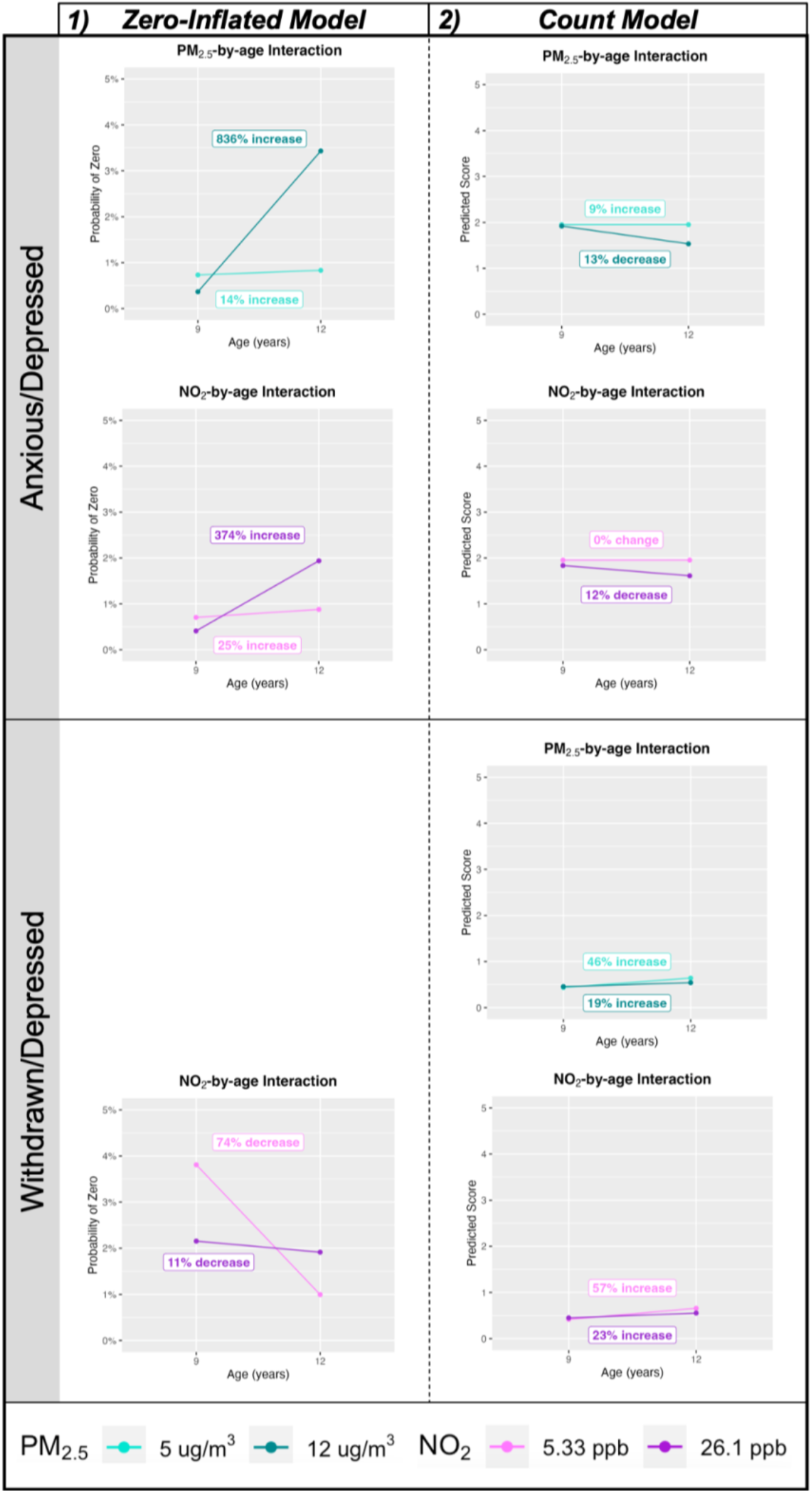
Significant results for anxious/depressed and withdrawn/depressed. NO_2_ is set to the WHO standard (5.33 ppd) for the PM_2.5_-by-age models and PM_2.5_ is set to the WHO standard (5 µg/m^3^) for the NO_2_-by-age models. ***Column 1)*** Displays the estimated probability of being in the absolute zero category as compared to the non-zero category (i.e., any value for CBCL scores). ***Column 2)*** Displays the estimated CBCL score for subjects whose scores were in the non-zero category. Numerous results are presented which include: ***a)* PM_2.5_-by-age interaction** which displays differences in 9 and 12 years- of-age for the WHO recommended PM_2.5_ levels - 5 µg/m^3^ (light blue) - versus the EPA’s - 12 µg/m^3^ (dark blue); ***b)* NO_2_-by- age interaction** which displays differences in 9 and 12 years-of-age for the WHO recommended NO_2_ levels - 5.33 ppd (light purple) - versus the 90^th^ percentile NO_2_ level in our sample - 26.1 ppd (dark purple) (The EPA level is 53 ppd which is outside our sample range). All graphs display percent change with age. All covariates held constant at the largest N category (sex = “male”, race/ethnicity = ‘White’, caregiver education = ‘Post Graduate Degree’, caregiver employment = “Employed”, and household income = “≥$100K”), and mean for neighborhood safety (𝑥 = 3.88).

#### 3.3.2 Rule-breaking and Aggressive Behavior

For rule-breaking and aggressive behavior, we found a significant main effect of age for the zero-inflated portion of the model, demonstrating a 112% and 106% increase in the probability of having zero rule-breaking and aggressive behavioral problems (i.e., obtaining a true-zero) with increasing age, respectively. For the count portion of the model, we found a significant main effect of age showing a 10% and 14% decrease in rule-breaking and aggressive behavioral problems, respectively, with increasing age (**Supplemental Figure 4)**. Similarly to externalizing problems, for rule-breaking and aggressive behavior there was no significant interaction between PM_2.5_ and age for either the zero-inflated or the count portions of the model (**Supplemental Table 5**). For the main effect of PM_2.5_ there was only a significant effect for aggressive behavior. Specifically, there was a similar trend that mirrored externalizing behavior, with a 63% increase in the probability of reporting no aggressive problems at a concentration of 12 µg/m^3^ as compared to 5 µg/m^3^. There was no significant main effect of PM_2.5_ in the count portion of either of these main effect models. For NO_2_, there was a significant interaction of NO_2_ and age in the zero-inflated portion of the model for rule-breaking behavior. At concentrations of 5.3 ppb there was a 27% increase in this probability of not having any rule-breaking problems from 9 to 12 years-of-age, whereas the probability of reporting no problems at a concentration of 26.1 ppb was much greater (175%). For the non-zero count portion of the model, the NO_2_-by-age was also significant for both rule-breaking and aggressive behavior with very similar trends as seen with overall externalizing problems (**Figure 3**). Again, the magnitude of probabilities for reporting no problems and changes in predicted problem scores were minimal.

**Figure 3.**
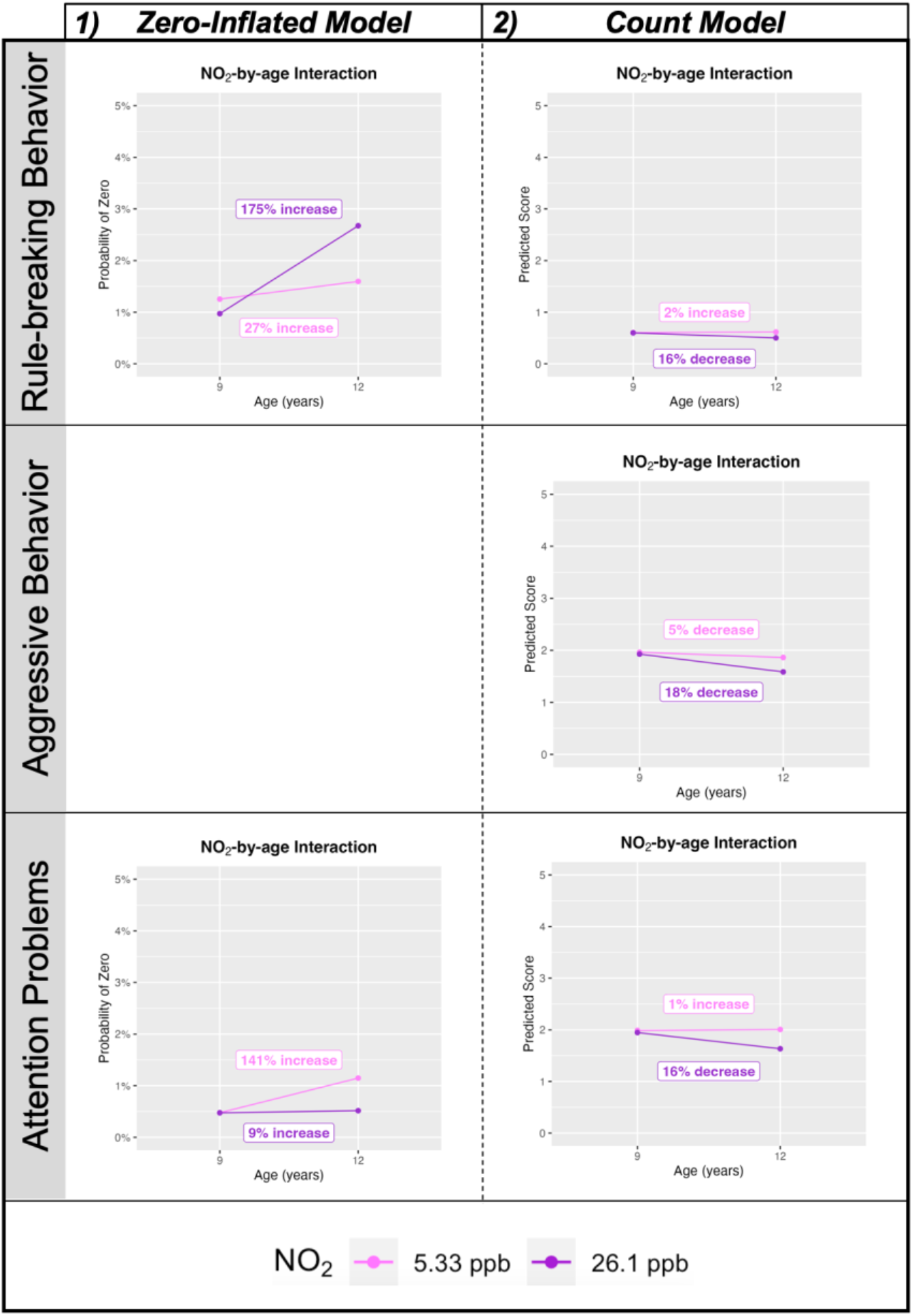
Significant results for rule-breaking behavior, aggressive behavior, and attention problems. NO_2_ is set to the WHO standard (5.33 ppd) for the PM_2.5_-by-age models and PM_2.5_ is set to the WHO standard (5 µg/m^3^) for the NO_2_-by-age models. ***Column 1)*** Displays the estimated probability of being in the absolute zero category as compared to the non-zero category (i.e., any value for CBCL scores). ***Column 2)*** Displays the estimated CBCL score for subjects whose scores were in the non-zero category. The only significant interactions seen for these subscales were the **NO_2_-by-age interaction** which displays differences in 9 and 12 years-of-age for the WHO recommended NO_2_ levels - 5.33 ppd (light purple) - versus the 90^th^ percentile NO_2_ level in our sample - 26.1 ppd (dark purple) (The EPA level is 53 ppd which is outside our sample range). All graphs display percent change with age. All covariates held constant at the largest N category (sex = “male”, race/ethnicity = ‘White’, caregiver education = ‘Post Graduate Degree’, caregiver employment = “Employed”, and household income = “≥$100K”), and mean for neighborhood safety (𝑥 = 3.88).

#### 3.3.3 Attention Problems

For attention problems, we found a significant main effect of age for the zero-inflated portion of the model, demonstrating a 58% increase in the probability of having zero externalizing problems (i.e., obtaining a true-zero) with increasing age, as well as for the count portion of the model, showing a 10% decrease in problems with increasing age (**Supplemental Figure 4)**. There was no significant interaction between PM_2.5_ and age for attention problems for either the zero-inflated or the count portions of the model. For the main effect of PM_2.5_, an 87% increase in the probability of reporting no attention problems (i.e., certain-zero) was seen at a PM_2.5_ concentration of 12 µg/m^3^ as compared to 5 µg/m^3^. Though, again, these absolute differences in probability are quite low. For the NO_2_-by-age model, we see a significant interaction in both the zero-inflated portion of the attention model and the non-zero count portion of the model (**Figure 3**). For the zero-inflated model, there is an opposite effect as compared to all the other models, with the lower concentration of 5.3 ppb of NO_2_ predicting the higher probability (141% increase) of having no attention problems (i.e., true-zero) from 9 to 12 years-of-age as compared to the concentration of 26.1 ppb. Though, as previously stated, this interaction still leads to a very small probability difference. For the non-zero count portion of the model, at the higher NO_2_ concentration of 26.1 ppb, less attention problems were seen from 9 to 12 years-of-age as compared to the low NO_2_ concentration of 5.3 ppb. However, the relative differences in change is less than a 1-point change in the attention problems score.

## Discussion

In the current longitudinal study, we leveraged a large, nationwide longitudinal cohort of children to examine how exposure to both PM_2.5_ and NO_2_ at ages 9-10 years is associated with behavioral problems as reported on the CBCL over a 2-year follow-up period. To characterize the developmental trajectory of behavioral problems within our sample as well as aid interpretation of the pollutant effects we examined the main effect of age on CBCL; in these models we saw that we observed an increased probability of zero problems (in the zero-inflated portion of the models) as well as fewer problems when problems were present (in the count portion of the models) in the internalizing, externalizing, anxious/depressed, rule-breaking, aggressive, and attention syndrome scales; interestingly, we saw the opposite effect all in the withdrawn/depressed syndrome scale, where increasing age was related to decreased probability of zero problems (in the zero-inflated portion of the models) as well as more problems when problems were present (in the count portion of the models). Though, in contrast to our hypothesis, incorporating each pollutant as a moderator of age led to higher levels of PM_2.5_ and NO_2_ exposure predicting a decreased likelihood of any internalizing problems and anxious/depressed behaviors with age as well as slightly fewer problems if these behaviors were present. Likewise, higher exposure to NO_2_ resulted in a decreased likelihood of any externalizing problems, including rule-breaking and aggressive problems with age, as well as less problems if present, whereas higher exposure to PM_2.5_ was also linked with a decreased likelihood of any externalizing and attention problems overall. In fact, only the association between NO_2_ exposure and presence of attention problems was in the expected direction, with lower NO_2_ exposure predicting an increased likelihood of an absence of attention problems as compared to those exposed to higher concentrations. While the directions of the relationships between the pollutants and CBCL outcomes are counterintuitive, it is important to consider the magnitude of the effect sizes in such a large sample, rather than the statistical significance of these findings. The effect sizes linking higher air pollution exposure to a smaller chance of any emotional or behavioral problems arising are ≤ 1.5% difference in probability. Moreover, the effect sizes are similarly small for quantitative differences in the number of problems, with higher pollution exposure associated with a decrease of less than a single point difference on any given scale. Given the 3-point likert scale of the CBCL (i.e., 0 = Not True, 1 = Somewhat or Sometimes True, 2 = Very True or Often True) used to endorse up to 118-items, a 1-point change is likely clinically negligible and may not have real-world implications. Thus, the small effect sizes seen in both the zero-inflated and count models indicate that this potential “protective” effect is likely to have very little, if any, clinical significance for onset of psychopathology in children and adolescents.

Our study focuses on childhood exposure at ages 9-10 years old – a developmental period currently underrepresented in the literature. About 26% of studies on pollution-related mental health problems cover this age range, despite the high incidence of psychiatric diagnoses in early adolescence (Kessler et al., 2005; Solmi et al., 2022; Zundel et al., 2022). Yet, even among these studies focused on linking PM_2.5_ and NO_2_ exposure and emotional behaviors in youth have reported mixed findings. Some of the earliest longitudinal research in this area comes from Columbia Center for Children’s Environmental Health (CCCEH) longitudinal cohort study of African-American and Dominican women in New York City. These essential studies found that prenatal exposure to airborne polycyclic aromatic hydrocarbons (PAHs), which come from fossil fuel combustion, was linked to greater CBCL reported symptoms of anxious/depressed and attention problems at ages 4-5 and 6-7 years-old children (Perera et al., 2011, 2012). However, in a more recent study that included using either the Strength and Difficulties Questionnaire or the CBCL in 8 European population-based birth cohorts, prenatal and postnatal air pollution, including PM_2.5_ and NO_2_ exposure, were not found to relate to the borderline clinical range of depression, anxiety, and aggression in >13,000 children ages 7-11 years-old (Jorcano et al., 2019). In fact, higher postnatal exposure was linked with overall lower odds of having symptoms in the borderline/clinical range when assessed cross-sectionally with the CBCL; albeit the results did not reach statistical significance. Similar findings were also noted when using the quantitative scores of the symptom scales (Jorcano et al., 2019) as implemented in the current analysis. A similar study assessing ADHD symptoms in children 3-10 years-old using these same 8 European birth cohorts also found no association, or even decreased risk, between prenatal air pollution exposure and ADHD (Forns et al., 2018). Thus, our current findings are in line with these more recent multi-research site-based studies. Similar to these studies, it seems very unlikely that the significant effects found in the current study are in fact reflective of a protective effect given both 1) the absence of any postulated mechanism for a protective element of air pollution exposure, as well as 2) the extremely small magnitude of change detected in part to our large sample size and the resulting statistical power. It is feasible that both the previous findings as well as the current results could be due to residual negative confounding (Forns et al., 2018; Jorcano et al., 2019), although it is important to note that in each case the analyses adjusted for many essential sociodemographic variables (i.e., income, caregiver educational attainment, etc.) that are known to be associated with air pollution exposure and mental health in children. Thus, if residual negative confounding is at play, that unexplained factor is likely to be pervasive across various cities within the western populations (e.g., U.S., Germany, Italy, Spain, etc.).

Putting our current results in the larger context of the literature, the importance of windows of exposure and the timing of behavior continue to prevail as to what role air pollution may play in terms of risk for mental health problems. That is, while the current study shows a one-year annual average of air pollution exposure during the transition to adolescence does not substantially increase the clinical risk of mental health problems over a 2-year follow-up period, it is still feasible that exposure during this period of development may ultimately predispose an individual to risk for psychopathology later in adolescence or early adulthood. Air pollution then may influence ongoing brain development and plasticity across adolescence, as regions and networks associated with mental health conditions and psychopathology (e.g., hippocampus, amygdala, default mode network, frontoparietal network, and salience network) (Menon, 2011, 2013) undergo protracted development. In fact, a number of MRI studies suggest that exposure to ambient air pollution is linked to differences in brain macro- and microarchitecture (Binter et al., 2022; Burnor et al., 2021; Essers et al., 2023; Guxens et al., 2018, 2022; Herting et al., 2019; Lubczyńska et al., 2021; Pérez-Crespo et al., 2022; Sukumaran et al., 2023). Thus, it is feasible that these differences may be early neural biomarkers of PM_2.5_ exposure-related risk prior to any overt changes in behavior. As previously mentioned, the idea that exposure during adolescence may ultimately predispose an individual to later develop mental health disorders parallel the findings that higher levels of air pollution during childhood and adolescence predicts later onset of major depressive disorder (Roberts et al., 2019) and other internalizing, externalizing, and thought disorder symptoms at age 18 years (Reuben et al., 2021). In fact, the increased incidence of psychopathology and psychiatric diagnoses seen in adolescence typically occurs in mid-adolescence, around age 14 and a half (Solmi et al., 2022), which is above the upper limit of ages included here. However, consortium efforts to eventually estimate lifetime air pollution exposure (C. C. Fan et al., 2021) in the coming years as well as active follow-up of ABCD cohort participants through early adulthood will ultimately allow for us to more formally test this hypothesis. Moreover, the results of the current study may also suggest that while PM_2.5_ and NO_2_ exposure at 9-10 years does not meaningfully impact the relative risk of emotional problems at a population-level, it is feasible that exposure during this time may have harmful effects in children who are more susceptible, due to either genetic risk or due to co-exposure to other adverse environmental threats. Thus, more research is warranted taking a more integrated neural exposome approach to understanding adolescent environmental exposures and risk for psychopathology (Tamiz et al., 2022).

The current study has a number of strengths. Specifically, the statistical approach and data used here contribute to a rigorous assessment of longitudinal behavioral and emotional problems associated with one-year annual air pollution exposure during the transition to adolescence. While standardized scores are often used to study dimensions of psychopathology and behavior between-subjects, we utilized raw longitudinal CBCL scores in the current study to better capture developmental change (Barch et al., 2021). However, raw CBCL scores are zero-inflated and over-dispersed in normative developmental samples, violating assumptions of general linear models. Our application of a zero-inflated negative binomial (ZINB) model combines the strengths of a logistic regression model with a negative binomial model, allowing robust estimates of how air pollution is related to the emergence of any behavioral or emotional problems (i.e., scores equal to zero vs. scores greater than zero), and how air pollution is related to the magnitude or number of behavioral or emotional problems (i.e., the range of scores greater than zero). Second, our large, nationwide sample between the ages of 9-12 years provides more geographically diverse estimates of NO_2_ and PM_2.5_. This is an improvement over the smaller, localized samples common to air pollution research, as sources and concentrations of pollutants vary across locations (Snider et al., 2016) and the health effects of PM_2.5_ vary by source (Holguin, 2008; Sarnat et al., 2008). Although the final sample used here is not fully representative of the larger US population (Garavan et al., 2018), it has greater generalizability as compared to other smaller scaled studies of air pollution and mental health and the models were adjusted for a large number of socioeconomic and lifestyle variables that are known to be associated with both exposure and emotional behaviors examined in the current study.

A limitation of the current study is that the estimates of air pollution used here only represent a sum across components of PM_2.5_ and capture a one-year annual average of exposure at the time of study enrollment. As previously mentioned, different geographical locations have different compositions of PM_2.5_ and the individual components of PM_2.5_ have different effects on human health. It is possible that our results represent an amalgamation of the unique effects of individual components of PM_2.5_ (e.g., elemental carbon, silicon, lead), contributing to our counterintuitive findings. There is also a substantial body of literature quantifying the effects of prenatal air pollution exposure as well as acute exposure (i.e., days) effects on various mental health outcomes (Braithwaite et al., 2019; Zundel et al., 2022), which are not available in the ABCD study at this time. Moving forward, incorporating prenatal exposure as well as acute estimates could help elucidate potential nuances that exist in the timing of exposure on the emergence of symptomatology across adolescence as well as the prevalence of acute mental health crises (for review, see (Heo et al., 2021). Another limitation is that the data included here were collected from 2016 until March 2020, at the beginning of the global COVID-19 pandemic. We chose to exclude data collected after March 2020, to avoid the confounding effect of pandemic-induced emotional and behavioral problems in this sample (Hamatani et al., 2022). The onset of the pandemic complicated data collection as well and may have contributed to missingness in data collected at later follow-up visits. For example, although sample demographics in the current study were similar to the larger ABCD cohort (**Supplemental Tables 1-3**) and our overall missingness was small (<=6%), we cannot rule out the possibility of selection bias influencing our results as not all participants had complete data at each wave of data collection, with follow-up waves having slightly higher representation of white children, with greater caregiver educational attainment and household income as compared to enrollment at baseline. However, due to this small proportion of missing data, that bias is expected to be small or negligible. Moreover, given that the ABCD study has under-representation of Asian, American Indian/Alaskan Native, and Native Hawaiian/Pacific Islander populations, as well as over-representation of families with higher total household incomes and highly educated caregivers, additional studies are needed that include children who may be especially susceptible to air pollution related effects because of potential compounding effects of disadvantage due to poverty and minority-related stressors stemming from racism (Hajat et al., 2015).

## Conclusions

The current study identified small negative associations between air pollution exposure and emotional behavior in children. The prevalence as well as the number of internalizing and externalizing symptoms were slightly lower over time for children exposed to higher levels of NO_2_ and PM_2.5_, showing that higher pollutant exposure somewhat exaggerated the main effects of age in most CBCL syndrome scales. Overall, these small effects are significant, but do not include changes that are likely to be clinically-relevant in terms of increased risk for psychopathology. Future research with additional waves of data extending into adolescence and incorporating cumulative exposure estimates and additional confounders are necessary to characterize links between air pollution and mental health during this period of heightened vulnerability during development.

## Credit authorship contribution statement

Claire E. Campbell: Project Administration, Methodology, Formal Analysis, Visualization, Writing - Original Draft; Devyn L. Cotter: Methodology, Writing - Original Draft; Katherine L. Bottenhorn: Methodology, Visualization, Writing – Original Draft; Elisabeth Burnor: Methodology, Data Curation, Writing – Review & Editing; Hedyeh Ahmadi: Data Curation, Methodology, Writing – Review & Editing; Carlos Cardenas-Iniguez: Writing - Review & Editing; W. James Gauderman: Methodology, Writing - Review & Editing; Rob McConnell: Methodology, Writing - Review & Editing; Kiros Berhane: Methodology, Writing - Review & Editing; Joel Schwartz: Methodology, Data Curation, Resources, Writing - Review & Editing; Jiu-Chiuan Chen: Conceptualization, Methodology, Writing - Review & Editing; Megan M. Herting: Funding Acquisition, Resources, Conceptualization, Methodology, Supervision, Project Administration, Writing - Original Draft.

## Data Availability

Data used in the preparation of this article were obtained from the Adolescent Brain Cognitive Development (ABCD) Study (https://abcdstudy.org), held in the NIMH Data Archive (NDA). This is a multisite, longitudinal study designed to recruit more than 10,000 children aged 9-10 and follow them over 10 years into early adulthood. ABCD consortium investigators designed and implemented the study and/or provided data but did not necessarily participate in analysis or writing of this report. This manuscript reflects the views of the authors and may not reflect the opinions or views of the NIH or ABCD consortium investigators. The ABCD data repository grows and changes over time. The ABCD data used in this report came from 10.15154/1523041.
R analyses code for this project can be found at 10.5281/zenodo.7787017.

https://zenodo.org/record/7787017#.ZEB0IezMKbs

https://nda.nih.gov/abcd/

## Acknowledgements

A special thank you to all ABCD cohort participants and their families. Portions of the research described in this article were conducted under contract to the HEI, an organization jointly funded by the US Environmental Protection Agency (EPA) and certain motor vehicle 14 and engine manufacturers. The content of this article does not necessarily reflect the views of HEI or its sponsors, nor does it necessarily reflect the views and policies of the US EPA or motor vehicle and engine manufacturers. Additional support for this research was also provided by the National Institutes of Health [NIEHS R01ES032295 (Herting), R01ES031074 (Herting), P30ES007048-23S1 (Gilliland), 3P30ES000002-55S1 (Weisskopf), NIHP01ES022845, T32 ES013678 (Campbell)] and EPA grants [RD-83587201 (Schwartz), RD-83544101 (Schwartz)] and the Rose Hills Foundation (MMH).

Data used in the preparation of this article were obtained from the Adolescent Brain Cognitive DevelopmentSM (ABCD) Study (https://abcdstudy.org), held in the NIMH Data Archive (NDA). This is a multisite, longitudinal study designed to recruit more than 10,000 children age 9-10 and follow them over 10 years into early adulthood. The ABCD Study® is supported by the National Institutes of Health and additional federal partners under award numbers U01DA041048, U01DA050989, U01DA051016, U01DA041022, U01DA051018, U01DA051037, U01DA050987, U01DA041174, U01DA041106, U01DA041117, U01DA041028, U01DA041134, U01DA050988, U01DA051039, U01DA041156, U01DA041025, U01DA041120, U01DA051038, U01DA041148, U01DA041093, U01DA041089, U24DA041123, U24DA041147. A full list of supporters is available at https://abcdstudy.org/federal-partners.html. A listing of participating sites and a complete listing of the study investigators can be found at https://abcdstudy.org/consortium_members/. ABCD consortium investigators designed and implemented the study and/or provided data but did not necessarily participate in the analysis or writing of this report. This manuscript reflects the views of the authors and may not reflect the opinions or views of the NIH or ABCD consortium investigators. Additional support for this work was made possible from NIEHS R01-ES032295 and R01-ES031074.

## Data and Code Availability Statement

Data used in the preparation of this article were obtained from the Adolescent Brain Cognitive Development (ABCD) Study (https://abcdstudy.org), held in the NIMH Data Archive (NDA). This is a multisite, longitudinal study designed to recruit more than 10,000 children aged 9-10 and follow them over 10 years into early adulthood. ABCD consortium investigators designed and implemented the study and/or provided data but did not necessarily participate in analysis or writing of this report. This manuscript reflects the views of the authors and may not reflect the opinions or views of the NIH or ABCD consortium investigators. The ABCD data repository grows and changes over time. The ABCD data used in this report came from <u>10.15154/1523041</u>.

R analyses code for this project can be found at <u>10.5281/zenodo.7787017</u>.

