## Supplemental Tables and Figures for "Air pollution and emotional behavior in adolescents across the U.S."

|  | Analytical Sample | Full ABCD Study |
| --- | --- | --- |
| Sex assigned at birth | | |
| Female | 4413 (47.6%) | 5658 (47.8%) |
| Male | 4858 (52.4%) | 6181 (52.2%) |
| Age at data collection | | |
| Mean (SD) | 118.856 (7.411) | 118.967 (7.495) |
| Range | 107.000 - 133.000 | 107.000 - 133.000 |
| Race/ethnicity | | |
| Missing values | 0 | 2 |
| Asian | 0 (0.0%) | 250 (2.1%) |
| Non-Hispanic Black | 1363 (14.7%) | 1777 (15.0%) |
| Hispanic | 1958 (21.1%) | 2405 (20.3%) |
| Non-Hispanic White | 4759 (51.3%) | 6162 (52.1%) |
| Other* | 1191 (12.8%) | 1243 (10.5%) |
| Caregiver education | | |
| Missing values | 0 | 14 |
| < HS Diploma | 460 (5.0%) | 592 (5.0%) |
| HS Diploma/GED | 899 (9.7%) | 1129 (9.5%) |
| Some College | 2423 (26.1%) | 3073 (26.0%) |
| Bachelor | 2310 (24.9%) | 3006 (25.4%) |
| Post Graduate Degree | 3179 (34.3%) | 4025 (34.0%) |
| Caregiver employment | | |
| Missing values | 0 | 56 |
| Employed | 6444 (69.5%) | 8194 (69.5%) |
| Stay at Home Parent | 1612 (17.4%) | 2065 (17.5%) |
| Unemployed | 539 (5.8%) | 669 (5.7%) |
| Other | 676 (7.3%) | 855 (7.3%) |
| Neighborhood safety | | |
| Missing values | 0 | 8 |
| Mean (SD) | 3.873 (0.976) | 3.890 (0.975) |
| Range | 1.000 - 5.000 | 1.000 - 5.000 |
| Household income | | |
| Missing values | 0 | 2 |
| <$50k | 2564 (27.7%) | 3215 (27.2%) |
| ≥$50K & <$100K | 2419 (26.1%) | 3065 (25.9%) |
| ≥$100K | 3514 (37.9%) | 4544 (38.4%) |
| Don't Know or Refuse | 774 (8.3%) | 1013 (8.6%) |

**Supplemental Table 1.** Baseline demographic comparison between the sample used here and the full ABCD Study sample. “Other” race/ethnicity category includes participants who were caregiver-identified as multi-racial, Asian, American Indian/Native American, Alaska Native, Native Hawaiian, Guamanian, Samoan, Other Pacific Islander, Asian Indian, Chinese, Filipino, Japanese, Korean, Vietnamese, Other Asian not listed, or Other Race not listed.

|  | Analytical Sample | Full ABCD Study |
| --- | --- | --- |
| Sex assigned at birth |  |  |
| Female | 4154 (47.4%) | 5335 (47.6%) |
| Male | 4605 (52.6%) | 5865 (52.4%) |
| Age at data collection |  |  |
| Mean (SD) | 130.924 (7.617) | 131.073 (7.714) |
| Range | 117.000 - 149.000 | 116.000 - 149.000 |
| Race, ethnicity |  |  |
| Missing Values | 0 | 2 |
| Asian | 0 (0.0%) | 239 (2.1%) |
| Non-Hispanic Black | 1221 (13.9%) | 1594 (14.2%) |
| Hispanic | 1800 (20.6%) | 2220 (19.8%) |
| Non-Hispanic White | 4612 (52.7%) | 5974 (53.3%) |
| Other | 1126 (12.9%) | 1171 (10.5%) |
| Caregiver education |  |  |
| Missing Values | 0 | 12 |
| < HS Diploma | 405 (4.6%) | 526 (4.7%) |
| HS Diploma/GED | 800 (9.1%) | 1007 (9.0%) |
| Some College | 2247 (25.7%) | 2847 (25.4%) |
| Bachelor | 2217 (25.3%) | 2889 (25.8%) |
| Post Graduate Degree | 3090 (35.3%) | 3919 (35.0%) |
| Caregiver employment |  |  |
| Missing Values | 0 | 47 |
| Employed | 6146 (70.2%) | 7826 (70.2%) |
| Stay at Home Parent | 1516 (17.3%) | 1941 (17.4%) |
| Unemployed | 481 (5.5%) | 595 (5.3%) |
| Other | 616 (7.0%) | 791 (7.1%) |
| Neighborhood safety |  |  |
| Missing Values | 0 | 5 |
| Mean (SD) | 3.884 (0.971) | 3.903 (0.969) |
| Range | 1.000 - 5.000 | 1.000 - 5.000 |
| Household income |  |  |
| Missing Values | 0 | 1 |
| <$50K | 2335 (26.7%) | 2930 (26.2%) |
| ≥$50K & <$100K | 2312 (26.4%) | 2937 (26.2%) |
| ≥$100K | 3418 (39.0%) | 4419 (39.5%) |
| Don't Know or Refuse | 694 (7.9%) | 913 (8.2%) |

**Supplemental Table 2**. Year one follow-up demographic comparison between the sample used here and the full ABCD Study sample. “Other” race/ethnicity category includes participants who were caregiver-identified as multi-racial, Asian, American Indian/Native American, Alaska Native, Native Hawaiian, Guamanian, Samoan, Other Pacific Islander, Asian Indian, Chinese, Filipino, Japanese, Korean, Vietnamese, Other Asian not listed, or Other Race not listed.

|  | Analytical Sample | Full ABCD Study |
| --- | --- | --- |
| Sex assigned at birth |  |  |
| Female | 2747 (47.1%) | 3481 (47.5%) |
| Male | 3080 (52.9%) | 3853 (52.5%) |
| Age at data collection |  |  |
| Mean (SD) | 143.081 (7.635) | 143.361 (7.747) |
| Range | 127.000 - 164.000 | 127.000 - 164.000 |
| Race, ethnicity |  |  |
| Missing Values | 0 | 0 |
| Asian | 0 (0.0%) | 158 (2.2%) |
| Non-Hispanic Black | 678 (11.6%) | 874 (11.9%) |
| Hispanic | 1198 (20.6%) | 1411 (19.2%) |
| Non-Hispanic White | 3236 (55.5%) | 4167 (56.8%) |
| Other | 715 (12.3%) | 724 (9.9%) |
| Caregiver education |  |  |
| Missing Values | 0 | 10 |
| < HS Diploma | 257 (4.4%) | 306 (4.2%) |
| HS Diploma/GED | 460 (7.9%) | 568 (7.8%) |
| Some College | 1466 (25.2%) | 1837 (25.1%) |
| Bachelor | 1551 (26.6%) | 2002 (27.3%) |
| Post Graduate Degree | 2093 (35.9%) | 2611 (35.6%) |
| Caregiver employment |  |  |
| Missing Values | 0 | 22 |
| Employed | 4139 (71.0%) | 5214 (71.3%) |
| Stay at Home Parent | 1002 (17.2%) | 1262 (17.3%) |
| Unemployed | 299 (5.1%) | 362 (5.0%) |
| Other | 387 (6.6%) | 474 (6.5%) |
| Neighborhood safety |  |  |
| Missing Values | 0 | 3 |
| Mean (SD) | 3.915 (0.948) | 3.938 (0.942) |
| Range | 1.000 - 5.000 | 1.000 - 5.000 |
| Household income |  |  |
| <$50k | 1494 (25.6%) | 1833 (25.0%) |
| ≥$50K & <$100K | 1597 (27.4%) | 2000 (27.3%) |
| ≥$100K | 2307 (39.6%) | 2952 (40.3%) |
| Don't Know or Refuse | 429 (7.4%) | 549 (7.5%) |

**Supplemental Table 3.** Year two follow-up demographic comparison between the sample used here and the full ABCD Study sample. “Other” race/ethnicity category includes participants who were caregiver-identified as multi-racial, Asian, American Indian/Native American, Alaska Native, Native Hawaiian, Guamanian, Samoan, Other Pacific Islander, Asian Indian, Chinese, Filipino, Japanese, Korean, Vietnamese, Other Asian not listed, or Other Race not listed.

|  | Baseline | 1-year follow-up | 2-year follow-up |
| --- | --- | --- | --- |
| Internalizing |  |  |  |
| Mean (SD) | 5.154 (5.539) | 5.281 (5.612) | 5.047 (5.610) |
| Range | 0.000 - 51.000 | 0.000 - 48.000 | 0.000 - 50.000 |
| Externalizing |  |  |  |
| Mean (SD) | 4.484 (5.798) | 4.232 (5.622) | 3.949 (5.368) |
| Range | 0.000 - 49.000 | 0.000 - 46.000 | 0.000 - 46.000 |
| Anxious/Depressed |  |  |  |
| Mean (SD) | 2.569 (3.060) | 2.613 (3.091) | 2.352 (2.941) |
| Range | 0.000 - 26.000 | 0.000 - 22.000 | 0.000 - 22.000 |
| Withdrawn/Depressed | |  |  |
| Mean (SD) | 1.045 (1.700) | 1.151 (1.800) | 1.235 (1.912) |
| Range | 0.000 - 14.000 | 0.000 - 14.000 | 0.000 - 16.000 |
| Rule-Breaking |  |  |  |
| Mean (SD) | 1.204 (1.844) | 1.142 (1.823) | 1.068 (1.829) |
| Range | 0.000 - 20.000 | 0.000 - 20.000 | 0.000 - 23.000 |
| Aggressive |  |  |  |
| Mean (SD) | 3.280 (4.305) | 3.090 (4.154) | 2.881 (3.894) |
| Range | 0.000 - 36.000 | 0.000 - 33.000 | 0.000 - 32.000 |
| Attention |  |  |  |
| Mean (SD) | 3.042 (3.508) | 2.941 (3.458) | 2.782 (3.324) |
| Range | 0.000 - 19.000 | 0.000 - 19.000 | 0.000 - 19.000 |

**Supplemental Table 4.** Behavioral descriptives from the Child Behavior Checklist (CBCL) questionnaire of the final sample across three waves of data collection.

| Coefficient (95% CI) | | |
| --- | --- | --- |
|  | Zero-inflated model | Count model |
| Internalizing |  |  |
| Age | -0.049 (-0.141, 0.043) | 0.028 (0.01, 0.045) |
| PM_2.5_ | -0.028 (-0.099, 0.043) | 0.008 (-0.014, 0.030) |
| Age*PM_2.5_ | **0.059 (0.032, 0.087)** | **-0.009 (-0.015, -0.004)** |
| Externalizing |  |  |
| Age | 0.201 (0.127, 0.275) | -0.024 (-0.043, -0.005) |
| PM_2.5_ | 0.055 (-0.005, 0.115) | -0.008 (-0.034, 0.019) |
| Age*PM_2.5_ | 0.005 (-0.018, 0.028) | -0.005 (-0.011, 0.001) |
| Anxious/Depressed |  |  |
| Age | 0.043 (-0.034, 0.121) | 0.001 (-0.019, 0.020) |
| PM_2.5_ | -0.099 (-0.162, -0.037) | -0.003 (-0.028, 0.022) |
| Age*PM_2.5_ | **0.102 (0.079, 0.124)** | **-0.009 (-0.016, -0.003)** |
| Withdrawn/Depressed |  |  |
| Age | -0.161 (-0.201, -0.121) | 0.123 (0.097, 0.149) |
| PM_2.5_ | 0.091 (0.059, 0.122) | 0.009 (-0.022, 0.039) |
| Age*PM_2.5_ | -0.014 (-0.027, -0.001) | **-0.011 (-0.019, -0.003)** |
| Rule-Breaking |  |  |
| Age | **0.234 (0.196, 0.272)** | -0.011 (-0.037, 0.015) |
| PM_2.5_ | -0.031 (-0.066, 0.005) | -0.009 (-0.039, 0.022) |
| Age*PM_2.5_ | 0.01 (-0.003, 0.022) | -0.007 (-0.015, 0.001) |
| Aggressive |  |  |
| Age | 0.311 (0.242, 0.381) | -0.029 (-0.048, -0.009) |
| PM_2.5_ | 0.112 (0.055, 0.168) | -0.008 (-0.036, 0.020) |
| Age*PM_2.5_ | -0.024 (-0.046, -0.003) | -0.006 (-0.012, 0.001) |
| Attention |  |  |
| Age | 0.208 (0.123, 0.293) | -0.028 (-0.047, -0.010) |
| PM_2.5_ | 0.123 (0.058, 0.189) | -0.005 (-0.032, 0.023) |
| Age*PM_2.5_ | -0.02 (-0.045, 0.005) | -0.003 (-0.008, 0.003) |

**Supplemental Table 5.** Model output for coefficients of interest for Age*PM_2.5_ interaction models. Bold values indicate coefficients of interest that passed the FDR test for multiple comparisons.

| Coefficient (95% CI) | | |
| --- | --- | --- |
|  | Zero-inflated model | Count model |
| Externalizing |  |  |
| Age | **0.215 (0.177, 0.253)** | **-0.039 (-0.048, -0.030)** |
| PM_2.5_ | **0.068 (0.028, 0.108)** | -0.017 (-0.041, 0.008) |
| Rule-Breaking |  |  |
| Age | **0.244 (0.223, 0.264)** | **-0.031 (-0.044, -0.019)** |
| PM_2.5_ | -0.003 (-0.027, 0.021) | -0.021 (-0.048, 0.007) |
| Aggressive |  |  |
| Age | **0.246 (0.210, 0.281)** | **-0.044 (-0.054, -0.034)** |
| PM_2.5_ | **0.070 (0.033, 0.108)** | -0.017 (-0.043, 0.009) |
| Attention |  |  |
| Age | **0.150 (0.109, 0.191)** | **-0.035 (-0.044, -0.026)** |
| PM_2.5_ | **0.090 (0.044, 0.136)** | -0.009 (-0.035, 0.017) |

**Supplemental Table 6.** Model output for coefficients of interest for Age+PM_2.5_ models. Bold values indicate coefficients of interest that passed the FDR test for multiple comparisons.

| Coefficient (95% CI) | | |
| --- | --- | --- |
|  | Zero-inflated model | Count model |
| Internalizing |  |  |
| Age | -0.042 (-0.155 - 0.071) | 0.040 (0.018 - 0.062) |
| NO_2_ | -0.03 (-0.05 - -0.01) | -0.001 (-0.001 - 0.005) |
| Age*NO_2_ | **0.013 (0.005 - 0.22)** | **-0.003 (-0.004 - -0.001)** |
| Externalizing |  |  |
| Age | 0.199 (0.104 - 0.294) | -0.006 (-0.03 - 0.018) |
| NO_2_ | -0.011 (-0.029 - 0.006) | -0.001 (-0.008 - 0.006) |
| Age*NO_2_ | 0.001 (-0.005 - 0.008) | **-0.002 (-0.004 - -0.001)** |
| Anxious/Depressed |  |  |
| Age | 0.074 (-0.024 - 0.172) | 0.001 (-0.025 - 0.026) |
| NO_2_ | -0.026 (-0.045 - -0.008) | -0.003 (-0.01 - 0.004) |
| Age*NO_2_ | **0.022 (0.015 - 0.029)** | **-0.002 (-0.004 - 0.0001)** |
| Withdrawn/Depressed |  |  |
| Age | -0.457 (-0.509 - -0.405) | 0.141 (0.108 - 0.174) |
| NO_2_ | -0.028 (-0.037 - -0.02) | 0.003 (-0.006 - 0.011) |
| Age*NO_2_ | **0.02 (0.016 - 0.024)** | **-0.004 (-0.006 - -0.001)** |
| Rule-Breaking |  |  |
| Age | 0.082 (0.031 - 0.132) | 0.007 (-0.025 - 0.04) |
| NO_2_ | -0.012 (-0.022 - -0.003) | -0.001 (-0.009 - 0.008) |
| Age*NO_2_ | **0.013 (0.009 - 0.016)** | **-0.003 (-0.005 - -0.001)** |
| Aggressive |  |  |
| Age | 0.262 (0.173 - 0.35) | -0.013 (-0.038 - 0.012) |
| NO_2_ | -0.003 (-0.02 - 0.013) | -0.001 (-0.008 - 0.007) |
| Age*NO_2_ | -0.001 (-0.007 - 0.005) | **-0.002 (-0.004 - -0.001)** |
| Attention |  |  |
| Age | 0.295 (0.196 - 0.395) | 0.005 (-0.018 - 0.029) |
| NO_2_ | -0.00007 (-0.018 - 0.018) | -0.001 (-0.008 - 0.006) |
| Age*NO_2_ | **-0.013 (-0.02 - -0.006)** | **-0.003 (-0.005 - -0.001)** |

**Supplemental Table 7.** Model output for coefficients of interest for Age*NO_2_ interaction models. Bold values indicate coefficients of interest that passed the FDR test for multiple comparisons.

| Variable Name in Manuscript Study | Variable Name/s from ABCD NDA | Description of ABCD NDA Variable | Structure Title | Structure Short Name |
| --- | --- | --- | --- | --- |
| subjectid | src_subject_id | Subject ID how it's defined in lab/project | ABCD Longitudinal Tracking | abcd_lt01 |
| eventname | eventname | The event name for which the data was collected | ABCD Longitudinal Tracking | abcd_lt01 |
| abcd_site | site_id_l | Site ID at each event | ABCD Longitudinal Tracking | abcd_lt01 |
| sex.bl | sex | Sex of subject at birth | ABCD Longitudinal Tracking | abcd_lt01 |
| interview_age | interview_age | Age in months at the time of the interview/test/sampling/imaging. | ABCD Longitudinal Tracking | abcd_lt01 |
| race_ethnicity.bl | *Recoded to include ‘Asian’ in ‘Other’ category given the low N* | | | |
|  | race_ethnicity | Race/Ethnicity (Child) | ABCD ACS Post Stratification Weights | acspsw03 |
| prnt.empl.bl | *Recoding as the following: employed = 1; unemployed = 2, 3, & 11; Stay at Home Parent = 6; Other = 4, 5, 7, 8, 9, 10; NA = 777* | | | |
|  | demo_prnt_empl_v2 | Caregiver employment status (baseline) | ABCD Parent Demographics Survey | pdem01 |
|  | demo_prnt_empl_v2_l | Caregiver employment status (longitudinal) | ABCD Longitudinal Parent Demographics Survey | abcd_lpds01 |
| overall.income.bl | *Recoding as the following: [<50k] = 1, 2, 3, 4, 5, & 6; [>=50K & <100K] = 7 & 8; [>100k] = 9 & 10; [Don't Know or Refuse] = 999 & 777* | | | |
|  | demo_comb_income_v2 | Combined Family Income | ABCD Longitudinal Parent Demographics Survey | abcd_lpds01 |
| high.educ.bl | *Recoding as the following: < HS Diploma = 0, 1, 2, 3, 4, 5, 6, 7, 8, 9, 10, 11, & 12; HS Diploma/GED = 13 & 14; Some College = 15, 16 & 17; Bachelor = 18; Post Graduate Degree = 19, 20 & 21* | | | |
|  | *demo_prnt_ed_v2* | Caregivers highest level of education attained | ABCD Parent Demographics Survey | pdem02 |
| neighb_phenx_avg_p.bl | *Recoding as the following: neighb_phenx_avg_p.bl = (neighborhood1r_p + neighborhood2r_p + neighborhood3r_p) / (# of questions answered, not including NA's)* | | | |
|  | *neighborhood1r_p* | 1st question in Crime Survey Modified from PhenX | ABCD Parent Neighborhood Safety/Crime Survey Modified from PhenX (NSC) | abcd_pnsc01 |
|  | *neighborhood2r_p* | 2nd question in Crime Survey Modified from PhenX | ABCD Parent Neighborhood Safety/Crime Survey Modified from PhenX (NSC) | abcd_pnsc01 |
|  | *neighborhood3r_p* | 3rd question in Crime Survey Modified from PhenX | ABCD Parent Neighborhood Safety/Crime Survey Modified from PhenX (NSC) | abcd_pnsc01 |
| reshist_addr1_pm252016aa_bl | *reshist_addr1_pm252016aa* | Residential history derived - annual average of PM 2.5 in 2016 at primary residential address at 1x1km2 | Residential History Derived Scores | abcd_rhds01 |
| reshist_addr1_no2_2016_aavg_bl | *reshist_addr1_no2_2016_aavg* | Residential history derived - annual average of NO2 in 2016 at primary residential address at 1x1km2 | Residential History Derived Scores | abcd_rhds01 |
| cbcl_scr_syn_internal_r | *cbcl_scr_syn_internal_r* | Internal CBCL Syndrome Scale (raw score) | ABCD Parent Child Behavior Checklist Scores Aseba (CBCL) | abcd_cbcls01 |
| cbcl_scr_syn_external_r | *cbcl_scr_syn_external_r* | External CBCL Syndrome Scale (raw score) | ABCD Parent Child Behavior Checklist Scores Aseba (CBCL) | abcd_cbcls01 |
| cbcl_scr_syn_anxdep_r | *cbcl_scr_syn_anxdep_r* | AnxDep CBCL Syndrome Scale (t-score) | ABCD Parent Child Behavior Checklist Scores Aseba (CBCL) | abcd_cbcls01 |
| cbcl_scr_syn_withdep_r | *cbcl_scr_syn_withdep_r* | WithDep CBCL Syndrome Scale (raw score) | ABCD Parent Child Behavior Checklist Scores Aseba (CBCL) | abcd_cbcls01 |
| cbcl_scr_syn_rulebreak_r | *cbcl_scr_syn_rulebreak_r* | RuleBreak CBCL Syndrome Scale (raw score) | ABCD Parent Child Behavior Checklist Scores Aseba (CBCL) | abcd_cbcls01 |
| cbcl_scr_syn_aggressive_r | *cbcl_scr_syn_aggressive_r* | Aggressive CBCL Syndrome Scale (raw score) | ABCD Parent Child Behavior Checklist Scores Aseba (CBCL) | abcd_cbcls01 |
| cbcl_scr_syn_attention_r | *cbcl_scr_syn_attention_r* | Attention CBCL Syndrome Scale (raw score) | ABCD Parent Child Behavior Checklist Scores Aseba (CBCL) | abcd_cbcls01 |

**Supplemental Table 8.** All variables used and their corresponding variable name within the ABCD Study downloadable database**.**

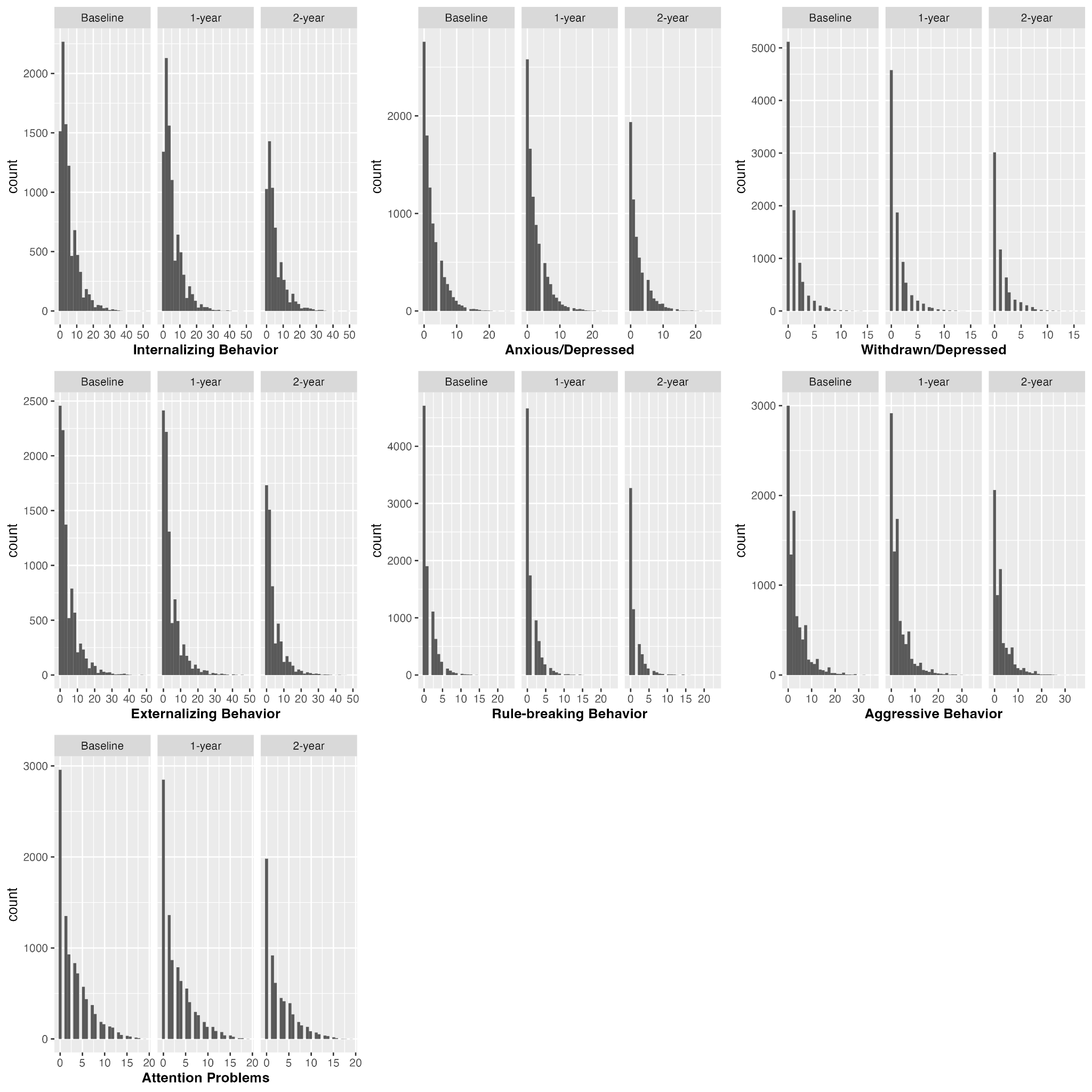

**Supplemental Figure 1** Histograms of each CBCL subscale score by collection wave (Baseline, 1-year, 2-year).

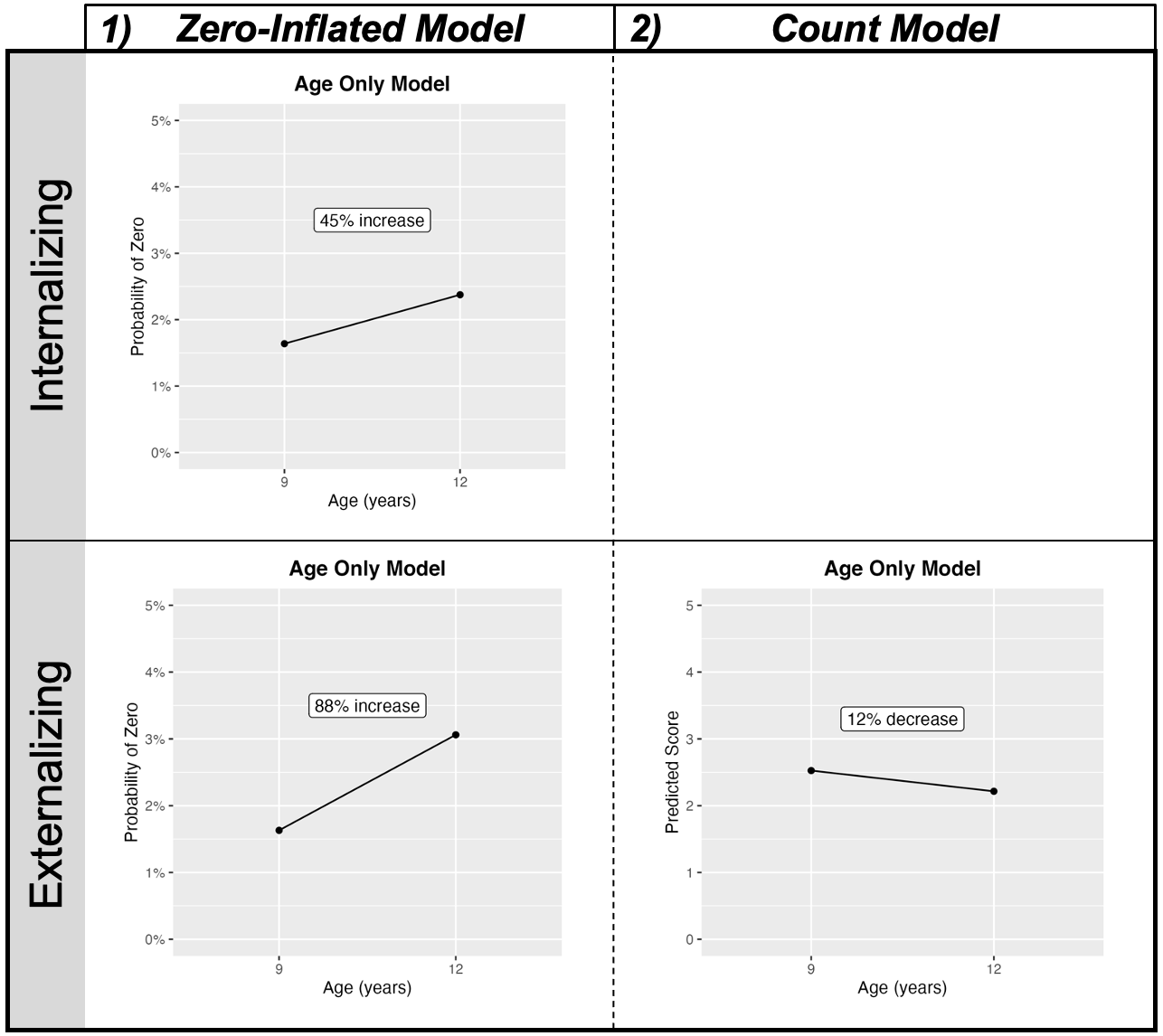

**Supplemental Figure 2** Results for significant main effects of age excluding air pollution with all other variables held constant on internalizing and externalizing behavior. The graphs display the estimated probability of being in the absolute zero category as compared to the non-zero category (i.e., any value for CBCL subscale scores) at age 9 versus 12 years-of-age. For all graphs, percent changes for age are displayed from 9 to 12 years-of-age. All covariates held constant at the largest N category (sex = “male”, race/ethnicity = ‘White’, caregiver education = ‘Post Graduate Degree’, caregiver employment = “Employed”, and household income = “≥$100K”), and mean for neighborhood safety ($\overline{x}$= 3.88).

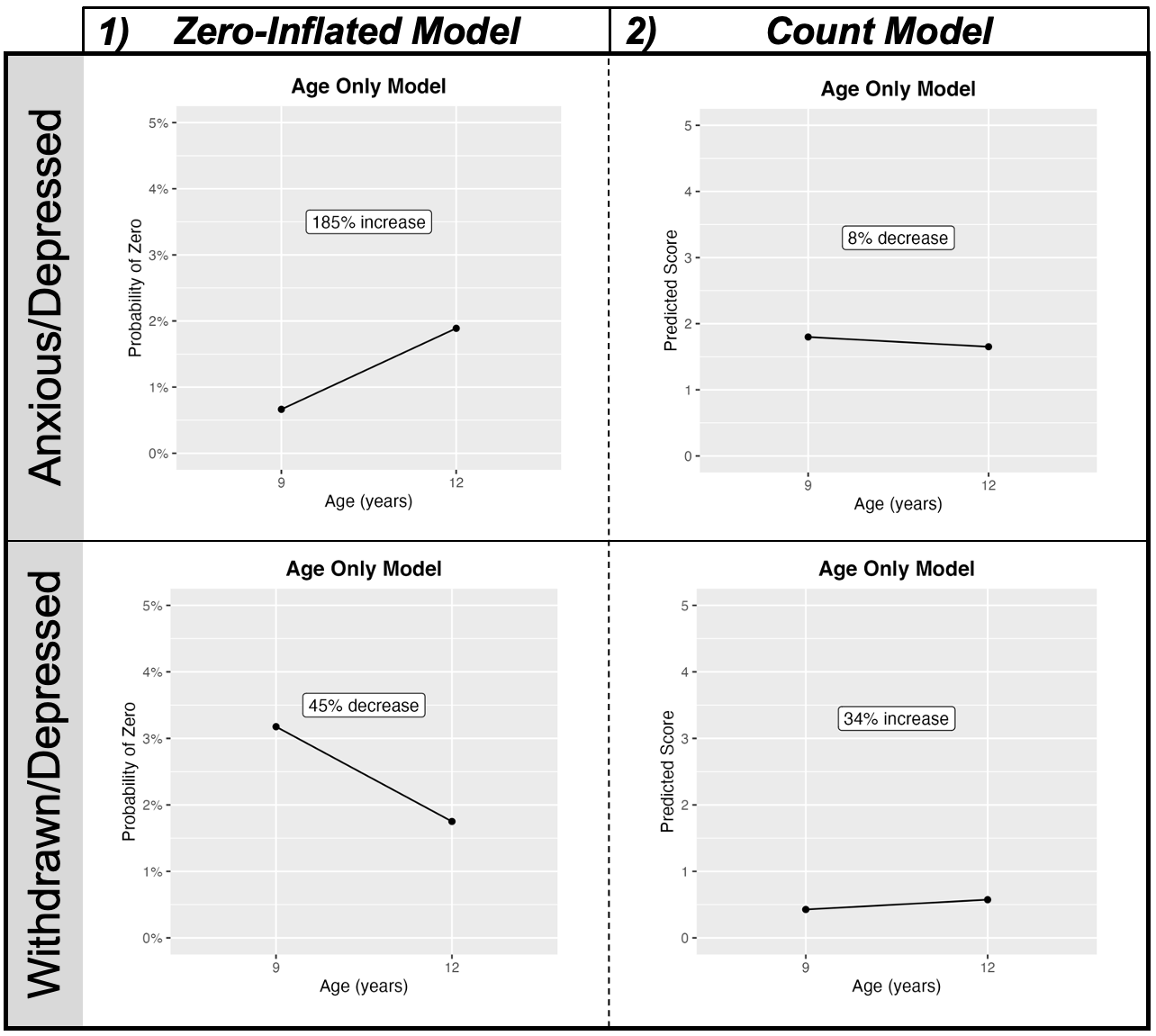

**Supplemental Figure 3** Results for significant main effects of age excluding air pollution with all other variables held constant on anxious/depressed and withdrawn/depressed. The graphs display the estimated probability of being in the absolute zero category as compared to the non-zero category (i.e., any value for CBCL subscale scores) at age 9 versus 12 years-of-age. For all graphs, percent changes for age are displayed from 9 to 12 years-of-age. All covariates held constant at the largest N category (sex = “male”, race/ethnicity = ‘White’, caregiver education = ‘Post Graduate Degree’, caregiver employment = “Employed”, and household income = “≥$100K”), and mean for neighborhood safety ($\overline{x}$= 3.88).

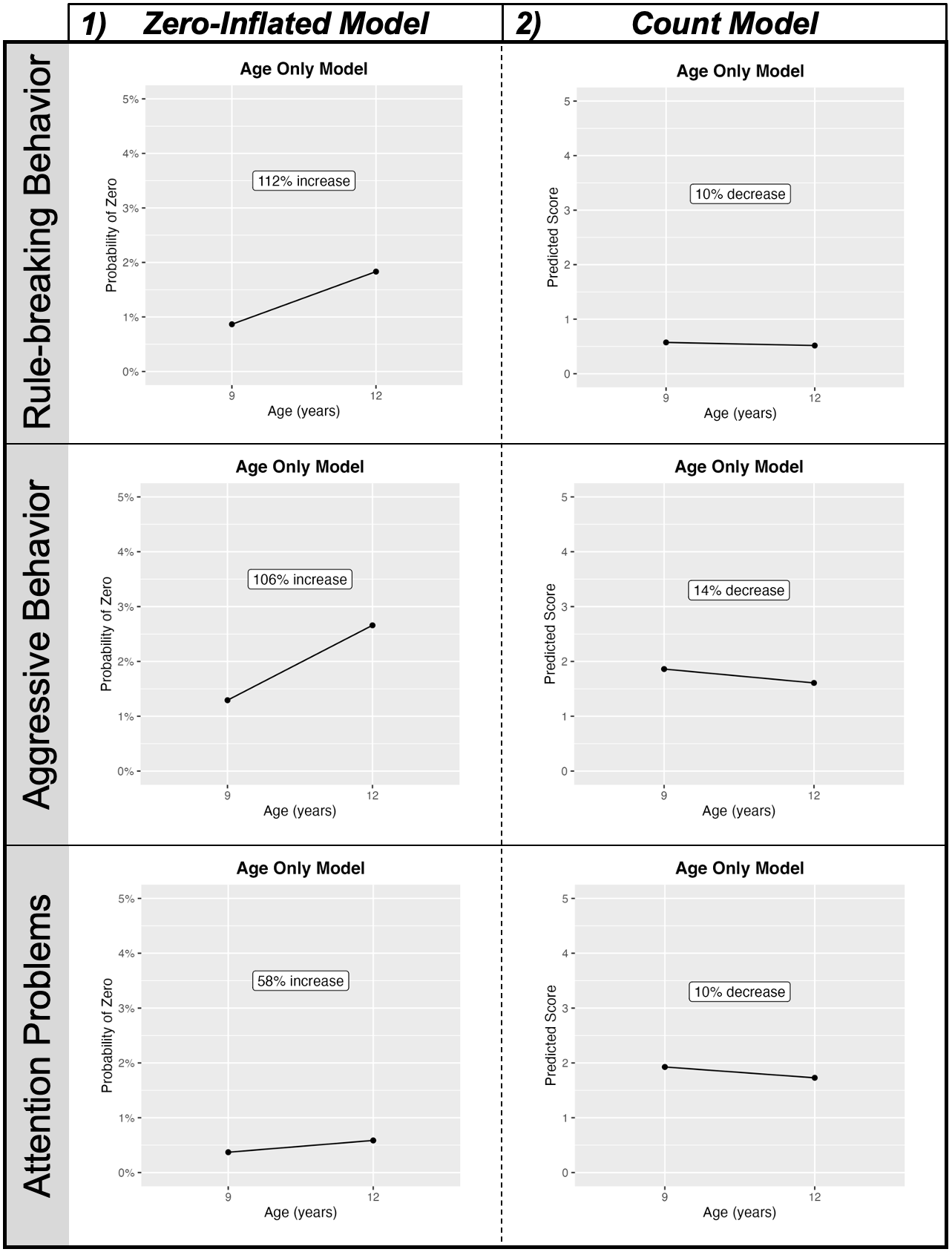

**Supplemental Figure 4** Results for significant main effects of age excluding air pollution with all other variables held constant on rule-breaking behavior, aggressive behavior, and attention problems. The graphs display the estimated probability of being in the absolute zero category as compared to the non-zero category (i.e., any value for CBCL subscale scores) at age 9 versus 12 years-of-age. For all graphs, percent changes for age are displayed from 9 to 12 years-of-age. All covariates held constant at the largest N category (sex = “male”, race/ethnicity = ‘White’, caregiver education = ‘Post Graduate Degree’, caregiver employment = “Employed”, and household income = “≥$100K”), mean for neighborhood safety ($\overline{x}$ = 3.88), and the WHO standards (PM_2.5_ = 5 µg/m^3^; NO_2_ = 5.33 ppd).

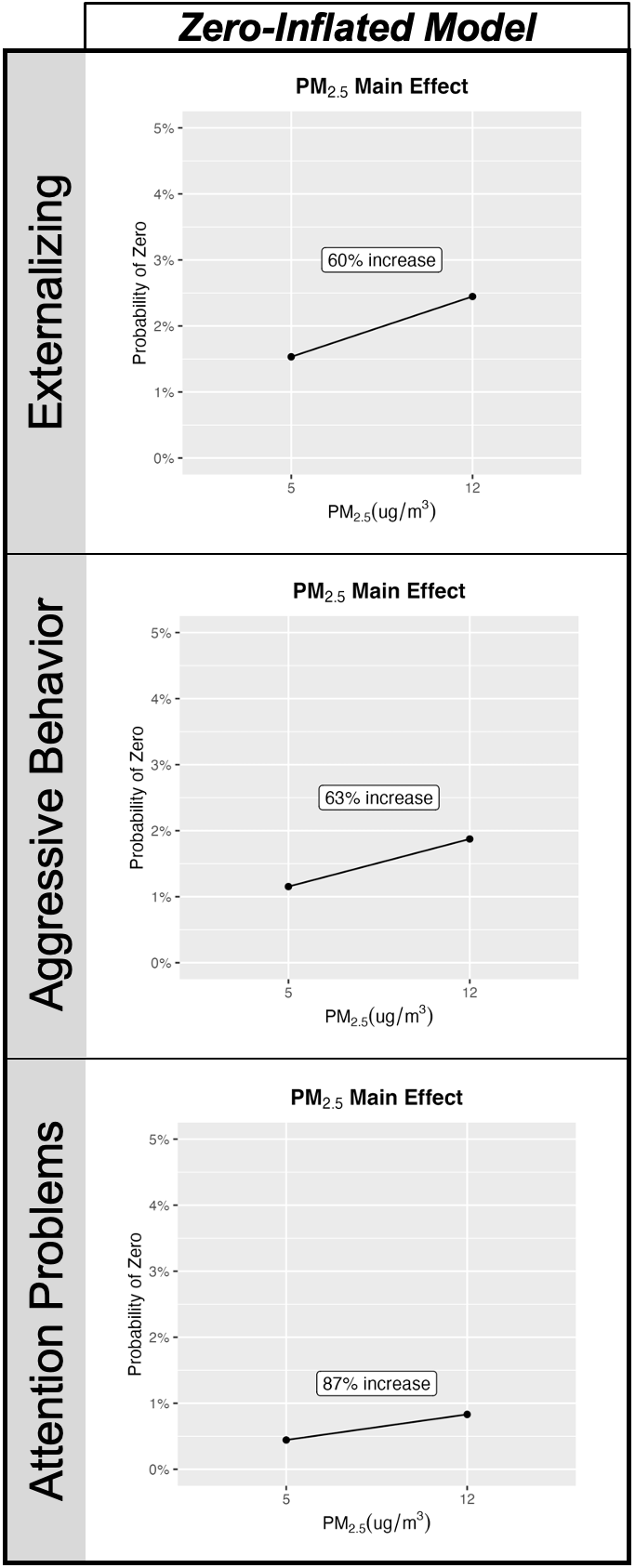

**Supplemental Figure 5** Significant results for the PM_2.5_ main effect. NO_2_ is set to the WHO standard (5.33 ppd) for the PM_2.5_ models. The column displays the estimated probability of being in the absolute zero category as compared to the non-zero category (i.e., any value for CBCL scores). The **PM_2.5_ main effect** displays the effect of pollutant at previously stated WHO versus EPA concentrations. All graphs display percent change with the increase in PM_2.5_. All covariates held constant at the largest N category (sex = “male”, race/ethnicity = ‘White’, caregiver education = ‘Post Graduate Degree’, caregiver employment = “Employed”, and household income = “≥$100K”), mean for neighborhood safety ($\overline{x}$= 3.88), and 9 years of age.
